# Large-Scale Whole-Exome Sequencing Association Study Implicates Genetic Effects on Viral Oncogenesis and Tumor Microenvironment in Nasopharyngeal Carcinoma

**DOI:** 10.1101/2023.10.18.23297131

**Authors:** Yanni Zeng, Chun-Ling Luo, Guo-Wang Lin, Fugui Li, Xiaomeng Bai, Josephine Mun-Yee Ko, Yang Liu, Shuai He, Jia-Xin Jiang, Wen-Xin Yan, Enya Hui Wen ONG, Zheng Li, Ya-Qing Zhou, Yun-He Zhou, An-Yi Xu, Shu-Qiang Liu, Yun-Miao Guo, Jie-Rong Chen, Xi-Xi Cheng, Yu-Lu Cao, Xia Yu, Biaohua Wu, Pan-Pan Wei, Zhao-Hui Ruan, Qiu-Yan Chen, Lin-Quan Tang, James D. McKay, Wei-Hua Jia, Hai-Qiang Mai, Jian-Jun Liu, Dong-Xin Lin, Chiea Chuen Khor, Melvin Lee Kiang CHUA, Mingfang Ji, Maria Li Lung, Yi-Xin Zeng, Jin-Xin Bei

## Abstract

Nasopharyngeal carcinoma (NPC) poses a substantial clinical challenge with limited understanding of its genetic underpinnings. Here we conduct the largest-scale whole-exome sequencing association study of NPC to date, involving 6,969 NPC cases and 7,100 controls and revealing three novel germline genetic variants linked to NPC susceptibility: a common variant rs2276868 in *RPL14*, a rare variant rs5361 in *SELE*, and a common variant rs1050462 in *HLA-B*. Through a multiomics approach, which integrates both bulk (n=206) and single-cell RNA-sequencing (n=56) data along with experimental validations, we demonstrate that the *RPL14* variant modulates Epstein-Barr virus (EBV) life cycle and NPC pathogenesis. Additionally, we show that the *SELE* variant plays a role in modifying endothelial cell function, thus promoting NPC progression. Our study also underscores the critical impact of rare genetic variants on NPC heritability. We introduce a refined composite polygenic risk score (rcPRS) that outperforms existing models in predicting NPC risk. Notably, our findings reveal that the polygenic risk for NPC is mediated by EBV infection status. Overall, our study provides crucial insights into the intricate genetic architecture of NPC. It highlights the critical interplay between genetic variations and essential elements of the tumor microenvironment, such as EBV and endothelial cells, in predisposing to NPC. This work opens new avenues for personalized risk assessments, early diagnosis, and targeted therapeutic strategies for NPC.

## Introduction

Nasopharyngeal carcinoma (NPC) is a highly lethal malignancy that exhibits a significant geographic prevalence in specific regions of East and Southeast Asia^1^. These areas account for approximately 70% of new cases worldwide^1^, with the highest incidence rate (9.69/100,000) reported in southern China area^2^. Familial clustering has been documented in a notable proportion (3.64∼19%) of NPC patients from diverse populations^3–6^, suggesting a strong link between family history and NPC risk^7^. Genetic susceptibility, Epstein-Barr virus (EBV) infection, and environmental effects are believed to play essential roles in NPC development^1^.

Efforts have been made to identify germline variations contributing to the genetic susceptibility of NPC. Multiple linkage and association studies have reported that common genetic variants in HLA region confer a significant risk for NPC^8–11^. Besides the HLA loci, linkage studies have also identified risk loci in 4p12-p15, 3p21.31-21.2, and 5p13.1^9,12,13^. However, the replication of these linkage results has been challenging due to the diminishing size of family pedigrees and the low resolution of reported loci, limiting their practical application. Recently, genome-wide association studies (GWASs) have identified replicated common single nucleotide polymorphism (SNPs) in multiple loci associated with NPC risk, including *MECOM*, *CDKN2A/B*, *TNFRSF19*, *CIITA*, and *TERT/CLPTM1L*, in addition to the HLA loci^8,14–17^. However, these GWAS loci explain only a small portion (2.84%) of the overall NPC susceptibility^18^, despite the relatively high heritability of NPC (10∼61.3%)^19,20^.

The issue of “missing heritability” (the gap between the heritability explained by GWAS loci and the total heritability) in NPC, similar to other complex polygenic diseases, may be attributed to the limited statistical power of studies targeting common variants (minor allele frequency (MAF)>=0.01) and even poorer power of studies capturing rare variants (MAF<0.01), such as those reported by utilizing whole-genome sequencing (WGS) or whole-exome sequencing (WES) ^21,22^. Currently, only a few studies have examined the contribution of rare variants to NPC risk through WES, identifying NPC-associated genes such as *MST1R, RAD54L* and *POLN* ^23–26^. However, these findings either did not reach exome-wide statistical significance or have not been independently validated, likely due to limited sample sizes or genetic heterogeneity across population. Therefore, the genetic architecture involving both common and rare variants in NPC risk remains unclear.

The role of EBV molecules in the screening and diagnosis of NPC is significant, as evidenced by their presence in NPC tumor and peripheral blood^27,28^. This is particularly remarkable given that over 90% of the adult population is infected with EBV^29^. However, the exact mechanisms linking EBV to NPC, and the potential involvement of host genetic variations in those processes, are still enigmatic. EBV is known for its ability to transform B cells into lymphoblastoid cell lines and for encoding oncogenic genes such as LMP1 and BARFs^30^. In NPC, EBV infects epithelial cells and encodes viral genes related to NPC progression^31^. Furthermore, the interaction of EBV with malignant epithelial cells and various stromal cell types, each exhibiting unique gene expression profiles, fosters a complex tumor microenvironment (TME), leading to both intra- and inter-tumor heterogeneity in NPC^32,33^. Despite these insights, the influence of genetic variations on TME components, and how this contributes to NPC predisposition remains elusive.

This study aims to deepen our understanding of the genetic architecture of NPC and to translate genetic findings into insights about its pathological mechanisms. We first conducted a comprehensive two-stage association study, utilizing WES and capture-sequencing (Cap-seq) analyses across multiple levels—variant, gene, and pathway—involving 14,069 individuals of southern Chinese descent, including those from Guangdong, Hong Kong in China, and Singapore. Furthermore, we employed extensive bulk and single cell RNA sequencing analyses, along with experimental assays, to elucidate the functional mechanisms of the identified risk loci. Critically, we uncovered the profound impact of both common and rare genetic variants on cancer heritability, particularly how these genetic variations distinctly affect EBV-related pathogenesis and TME components, thereby contributing to varying susceptibilities to NPC.

## Results

### Association of single variants with NPC susceptibility

To identify susceptibility loci of NPC at single variant level, we first performed whole-exome sequencing (WES) on genomic DNA samples for the discovery stage, involving 2,694 NPC cases and 2,328 healthy controls of Han Chinese descent from southern China (GD-SYSUCC) and Singapore (SG; see Methods, **Figure 1**). After stringent quality controls, we obtained 1,043,522 single nucleotide variants (SNVs), including 362,993 potentially functional or pathogenic variants (Methods) and 335,163 novel variants which were not present in the 1000 genome phase 3, gnomAD exome, and genome database^34,35^.

**Figure 1.**
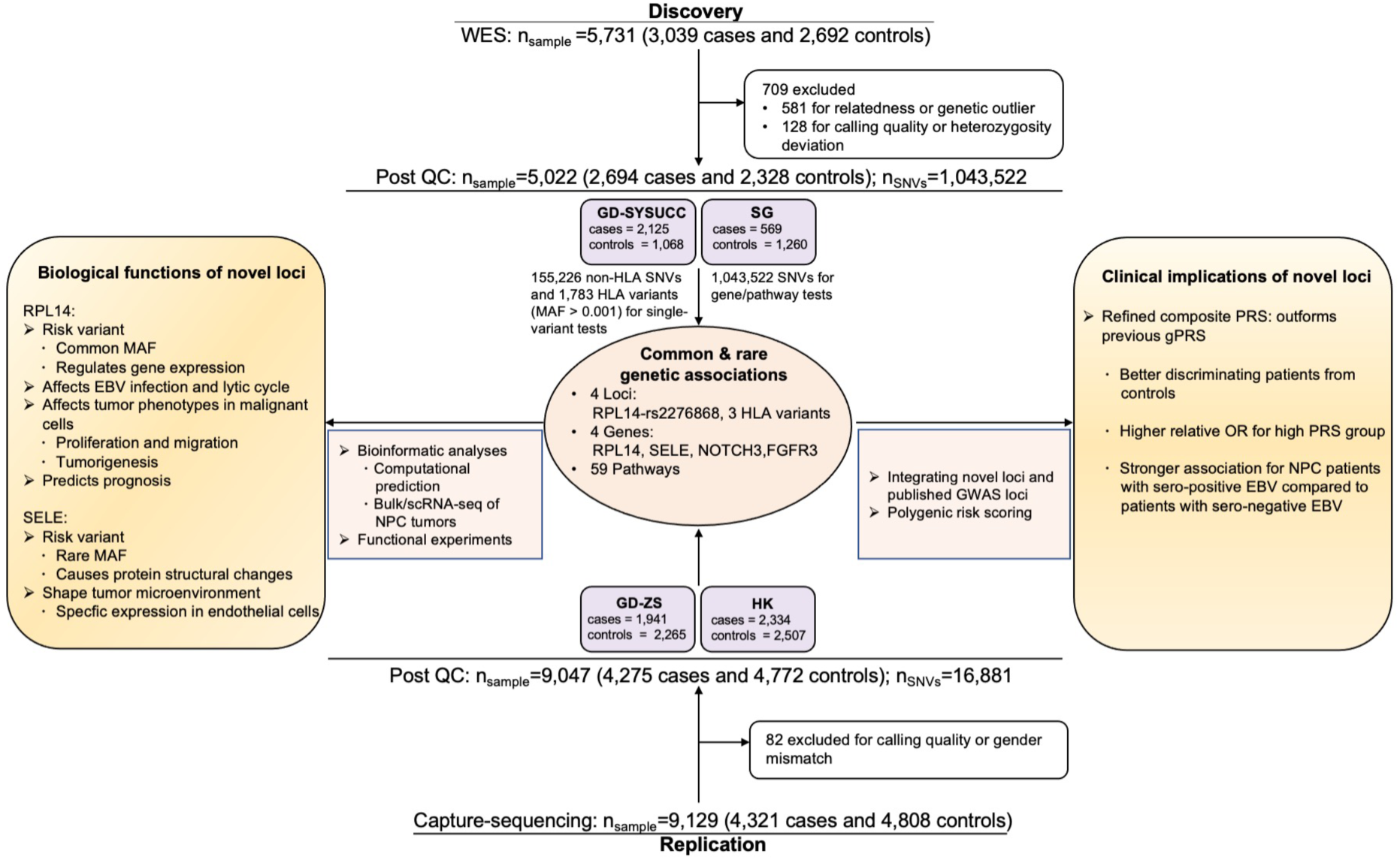
Study overview. A two-stage association study design was applied to investigate the genetic factors associated with NPC. In the discovery stage, a total of 5,022 samples, including the GD-SYSUCC cohort from Guangdong in China and the SG cohort from Singapore, were genotyped with WES and analyzed to identify four independent variants (two of which are novel), four novel genes, and 59 pathways associated with NPC. The associations were subsequently validated in the replication stage, which included 9,047 samples from the GD-ZS cohort in Guangdong and the HK cohort in Hong Kong, China. Bioinformatic analyses and functional experiments were conducted to explore the biological functions and the clinical implications of the novel loci.

Subsequently, we performed a single-variant-based exome-wide association study (EWAS) on 155,226 non-HLA SNVs and 1,783 HLA variants (including HLA alleles, amino acids [AA], and SNVs) with a MAF >= 0.001 among 2,694 NPC cases (all NPC cases, ALLNPC) and 2,328 controls, as well as 409 NPC cases with a family history (FHNPC) and the same set of controls. We observed neither evidence of genomic inflation of the associations (λ_ALLNPC_=1.07, λ_FHNPC_=0.89, **Figure S1A**) nor significant genetic substructure between case and control groups (**Figure S2**). The EWAS results revealed a total of 242 variants associated with NPC risk in the ALLNPC group and 128 variants associated with NPC risk in the FHNPC group, surpassing the Bonferroni significant threshold for multiple testing correction (P < 3.2 × 10^-7^; **Figure S1B**). Notably, we discovered a novel non-HLA locus with several significant associated SNVs (**Figure S1B**). The sentinel SNV rs2276868, located at the 5’-untranslated region (5’-UTR) of the ribosomal gene *RPL14*, exhibited the strongest associations in both the FHNPC (P_FHNPC_=2.3 × 10^-8^, Odds Ratio (OR) _FHNPC_ =1.575, 95% Confident Interval (CI) =1.343-1.846) and the ALLNPC groups (P_ALLNPC_=4.2 × 10^-7^, OR_ALLNPC_ =1.242, 95%CI=1.142-1.351; **Table 1, Figure S1B**). Furthermore, the association was stronger in the family cases than in the sporadic cases (OR_sporadic_NPC_ =1.200, 95%CI =1.097-1.313), indicating a robust genetic predisposition to NPC. Conditional analysis revealed that all associations at the *RPL14* locus vanished upon adjusting for rs2276868, suggesting that this variant explains all the observed associations (**Table S1, S2**). Beyond the non-HLA locus, we also identified HLA variants significantly associated with NPC (**Figure S1B**). Stepwise conditional analysis localized three independently associated variants, including *HLA-DQB1*03:01:01*, *HLA-A: A_62_Q*, and rs1050462 at HLA-B (**Tables 1, Table S1**). Except for the two known associations at *HLA-A: A_62_Q* and *HLA-DQB1*03:01:01*^18,36^, the association of rs1050462 with NPC is a novel discovery whose signal is not attributed to previously reported *HLA* variants ^8,10,11^ (see Methods; P ≤ 3.23 × 10^-5^; **Table S3**), according to an additional conditional analysis using an extended GD-SYSUCC cohort consisting of 1,583 cases and 3,049 controls with GWAS array data ^17^.

**Table 1.** Association of single variants with NPC susceptibility.

| Variants information |  |  |  | Discovery<br>(2,694 cases and 2,328 controls) |  |  |  | Replication<br>(4,275 cases and 4,772 controls) |  |  |  |  | Combined samples<br>(6,969 cases and 7,100 controls) |  |  |
| --- | --- | --- | --- | --- | --- | --- | --- | --- | --- | --- | --- | --- | --- | --- | --- |
| Variant ID | CHR | A1 | Gene | ALLNPC:<br>OR (95% CI) | P | FHNPC:<br>OR (95% CI) | P | OR (95% CI)<br>HK<br>(2,334 cases and 2,507 controls) | OR (95% CI)<br>GD-ZS<br>(1,941 cases and 2,265 controls) | OR<sub>meta</sub>:<br>(95% CI) | P<sub>meta</sub> | I<sub>meta</sub><sup>2</sup> | OR<sub>meta</sub><br>(95% CI) | P<sub>meta</sub> | I<sub>meta</sub><sup>2</sup> |
| rs2276868 | 3 | T | RPL14 | 1.242 (1.142-1.351) | 4.16E-07 | 1.575 (1.343-1.846) | 2.30E-08 | 1.253 (1.154-1.36) | 1.193 (1.091-1.305) | 1.225 (1.153-1.302) | 5.26E-11 | 0.00 | 1.231 (1.172-1.293) | 1.22E-16 | 0.00 |
| DQB1*03:01:01 | 6 | Present | HLA-DQB1 | 0.633 (0.568-0.705) | 1.00E-16 | 0.585 (0.467-0.734) | 3.64E-06 | 0.694 (0.622-0.774) | 0.589 (0.522-0.663) | 0.640 (0.545-0.752) | 5.56E-08 | 74.84 | 0.638 (0.582-0.700) | 9.43E-22 | 50.44 |
| A_62_Q | 6 | Present | HLA-A | 0.507 (0.462-0.556) | 4.61E-47 | 0.483 (0.401-0.581) | 1.20E-14 | 0.585 (0.532-0.642) | 0.560 (0.505-0.621) | 0.573 (0.535-0.615) | 1.73E-55 | 0.00 | 0.549 (0.504-0.598) | 4.69E-43 | 57.52 |
| rs1050462 | 6 | G | HLA-B | 0.646 (0.585-0.713) | 7.58E-18 | 0.637 (0.521-0.779) | 1.08E-05 | 0.568 (0.513-0.629) | 0.588 (0.526-0.658) | 0.577 (0.535-0.622) | 1.92E-46 | 0.00 | 0.600 (0.555-0.649) | 2.55E-37 | 40.86 |

Notes: CHR, chromosome; A1, target allele/amino acid; OR (95%CI), odds ratio estimate and the 95% confidence interval; I<sup>2</sup>, the proportion of the variance in observed effect is due to variance in true effects rather than sampling error; Meta, meta-analyzed using the random effects model.

We next conducted validations of the identified associations using two independent replication cohorts consisting of 9,047 samples of Southern Chinese recruited from Hong Kong (HK; 2,334 cases and 2,507 controls) and Guangdong (GD-ZS; 1,941 cases and 2,265 controls) in China (see Methods). We genotyped the samples using Cap-seq to target the genomic regions spanning the above variants. Meta-analysis across the replication samples consistently confirmed the associations of the four variants in *RPL14* and *HLA* loci (P_meta_ < 0.05; **Table 1**).

Moreover, when combining the discovery and replication samples, the meta-analysis revealed that the four variants in *RPL14* and *HLA* loci were significantly associated with NPC risk, surpassing the threshold for genome-wide significance (P< 5 × 10^-8^; **Table 1**).

### Association of genes and pathways with NPC susceptibility

To identify the cumulative genetic effects of both rare and common variants within individual genes on the risks of NPC, we conducted a gene-based association analysis in the discovery dataset using an ensemble method that incorporates four algorithms simultaneously (see Methods). For each gene, we performed separate tests that use all SNVs within the genic region or only consider coding-affecting SNVs of that gene. Our analysis revealed a significant association between *RPL14* and NPC, with statistical significance surpassing Bonferroni threshold for multiple testing correction (P < 2.3 × 10^-6^, **Table 2**). Additionally, we validated previous associations in several genes (**Table S4**).

**Table 2.** Associations of individual genes with NPC susceptibility.

| Gene | Discovery (2,694 cases and 2,328 controls) |  |  |  | Replication<br>(4,275 cases and 4,772 controls) |  |  |  | Combined samples<br>(6,969 cases and 7,100 controls) |
| --- | --- | --- | --- | --- | --- | --- | --- | --- | --- |
| Name | Significant in tests | SNV group | Method | P | P<br>(GD-ZS; 1,941 cases<br>and 2,265 controls) | P<br>(HK; 2,334 cases<br>and 2,507 controls) | P<sub>meta</sub> | Adjusted<br>P<sub>meta</sub> | P<sub>meta</sub> |
| RPL14 | Gene association | ALLSNV | SKAT (C+R) | 2.17E-06 | 3.50E-06 | 8.44E-05 | 1.23E-07 | 6.16E-07 | 1.79E-10 |
| SELE | Sentinel pathway genes | ALLSNV | SKAT (C+R) | 2.69E-06 | 1.01E-03 | 3.94E-02 | 4.05E-04 | 2.02E-03 | 1.44E-07 |
| SELE | Sentinel pathway genes | coding<br>affecting<br>SNV | SKAT (C+R) | 3.61E-06 | 1.36E-03 | 1.20E-01 | 1.40E-03 | 7.00E-03 | 4.63E-07 |
| FGFR3 | Sentinel pathway genes | ALLSNV | Burden | 1.24E-04 | 7.40E-04 | 1.28E-01 | 8.96E-04 | 4.48E-03 | 3.64E-06 |
| NOTCH3 | Sentinel pathway genes | ALLSNV | SKAT (C+R) | 3.60E-04 | 6.00E-06 | 1.07E-02 | 3.28E-06 | 1.64E-05 | 5.41E-08 |

Notes: ALLSNV, all SNVs; SKAT (C+R), SNP-set (Sequence) Kernel Association Test for the combined effect of common (C) and rare (R) variants; Burden, burden test; Meta, meta-analyzed using the summation of logits method; Adjusted P, the P value adjusted using the FDR method.

We further performed a pathway-based analysis to explore the cumulative genetic effects of genes involved in 6,204 curated molecular pathways in the discovery dataset (see Methods). This analysis identified a total of 59 pathways associated with NPC in either the ALLNPC or FHNPC groups, adhering to the false discovery rate (FDR) threshold set for multiple testing correction (P < 3.09 × 10^-4^; **Figure 2A, Table S5)**. Among these pathways, we observed that 11 sentinel pathway genes were involved in at least two NPC-associated pathways and exhibited an association with NPC in gene-based test with a significant threshold of P < 0.01 (**Figure 2B**). Notably, *SELE, NOTCH3*, and *FGFR3* had strong associations with NPC (P < 1×10^-4^; **Figure 2B**). *SELE* was particularly outstanding, with the strongest association with NPC (P_SELE_ = 2.69 × 10^-6^) and the involvement in the largest number of NPC-associated pathways (**Figure 2B**).

**Figure 2.**
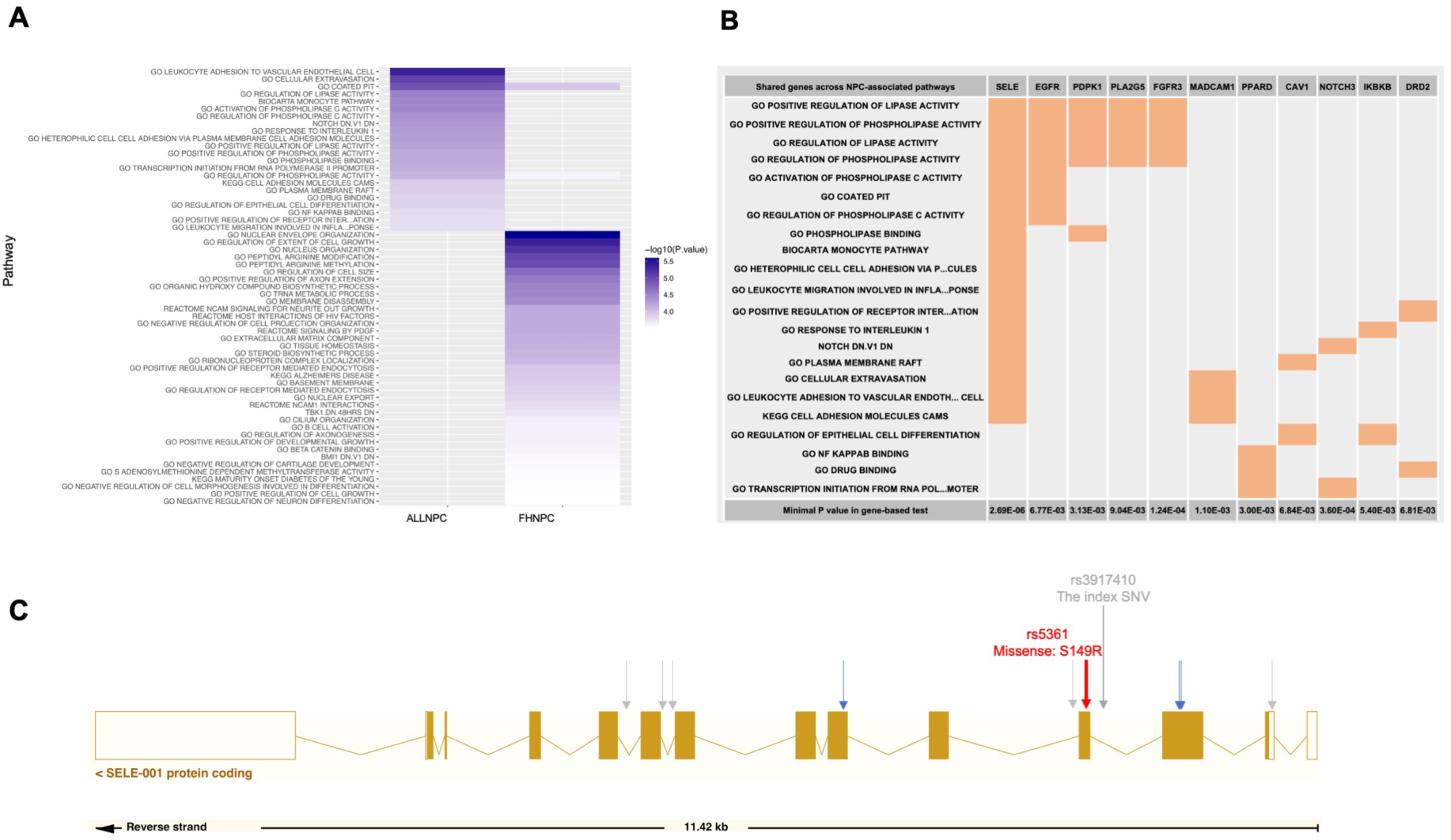
Significant pathways and sentinel genes associated with NPC. **A**. Significant pathways associated with NPC. ALLNPC denotes all NPC samples in the discovery stage, whereas FHNPC represents NPC cases with a familial NPC history. Color bars from white to purple indicate minus log-transformed P values. **B**. Sentinel genes for NPC-associated pathways. Genes indicated atop are highlighted in orange if they are part of a specific pathway. The listed genes are those belonging to at least two NPC-associated pathways and exhibiting a gene-based association P value below 0.01. **C**. Locations of the rare variants associated with NPC at significance level of P < 1×10^-4^ within the genic regions of SELE. Grey arrows denote non-coding variants, blue arrows represent synonymous coding variants, and the red arrow indicates non-synonymous coding variant predicted as “deleterious”. The rs5361 minor allele introduces a missense mutation at position 149, resulting in an amino acid substitution from S to R. The index SNV rs3917410 showed the most significant P value in the association tests in the discovery stage.

To validate the associations for the significant gene *RPL14* and the top three sentinel pathways genes (*SELE*, *FGFR3*, and *NOTCH3*), we re-sequenced exon regions of these four candidate genes in the replication cohorts (n_GD-ZS_=4,206 and n_HK_=4,841). Gene-based association analyses statistically validated all the candidate genes in the replication dataset (P_RPL14_=1.23 × 10^-7^, P_SELE_=4.05 × 10^-4^, P_NOTCH3_=3.28 × 10^-6^, and P*_FGFR3_*=8.96 × 10^-4^; **Table 2**).

The association for *RPL14* was particularly robust across all replication cohorts (P_GD-ZS_=3.50

× 10^-6^ and P_HK_=8.44 × 10^-5^), whereas the associations for *SELE*, *NOTCH3*, and *FGFR3* were more prominent in the GD-ZS cohort (**Table 2**).

We next fine-mapped the variants (MAF > 0.001) contributing to the gene-level associations for the four replicated genes using the discovery dataset (n=5,022). A constituent variant was considered to have a “major” contribution if its variant-level association P value was less than 1 × 10^-4^. Using this criterion, we detected a major contribution from a common variant for *RPL14* (**Table S6**), as well as major contributions from multiple rare variants for *SELE* (**Table S7**). However, no major contributing variants were detected for *FGFR3* (**Table S8**) or *NOTCH3* (**Table S9**). Specifically, among the four SNVs (MAF > 0.001) in *RPL14*, the sentinel SNV rs2276868 (MAF=0.36, P_SNV_=4.16 × 10^-7^) and another SNV rs2276869 (MAF=0.10, P_SNV_=0.001) shared modest linkage disequilibrium (LD; R^2^=0.2) and were the only two *RPL14* variants associated with NPC (P_SNV_ <0.05; **Table S6**). Removing these two SNVs from the gene-based analysis diminished the gene-level association of *RPL14* with NPC (P_gene_=0.34). Controlling for rs2276868 also abolished the associations of rs2276869 (**Table S6**). These findings suggest a predominant contribution of common variant rs2276868 to *RPL14*’s association with NPC.

For *SELE*, a total of 23 SNVs (MAF > 0.001) were detected, including seven common (MAF>=0.01) and 16 rare variants (MAF<0.01; **Table S7**). Of these, 10 SNVs were strongly associated with NPC (P_SNV_=2.68 × 10^-5^ ∼ 4.98 × 10^-5^; **Table S7**), with rs3917410 as the sentinel SNV (P=2.68 × 10^-5^) and sharing nearly complete LD with the other nine SNVs (R^2^ > 0.99; **Table S7**). All the 10 SNVs are rare, including one non-synonymous coding (rs5361), three synonymous coding, and six non-coding variants. Controlling for rs5361 variant abolished the association signals with NPC for the other rare variants in *SELE* (Adjusted P>0.05; **Table S7**). The rs5361 variant is a missense mutation (T>G) in the exon 4 of *SELE* (**Figure 2C**), resulting in an amino acid substitution from the conservative Serine (S) (**Figure 2C, Figure S3A**) to Arginine (R) at position 149. Furthermore, multiple algorithms predicted this S149R substitution as “deleterious” (Polyphen-2 score=0.997; CADD score=23; SIFT score=0; REVEL score=0.516; MetaLR score=0.772; **Figure S3B**). Collectively, these findings suggest that the rare variant rs5361 (*SELE*-S149R) is likely a causal variant accounting for the NPC associations observed in *SELE*.

### Functional characterization of the novel loci

#### Distinct cellular expression patterns of NPC-associated genes

To understand the functional consequences of the novel loci in NPC, we first investigated their cellular expression patterns along with the known NPC-associated genes (**Table S10**). We harnessed single-cell transcriptomic analyses of tumor samples from 56 NPC patients and various non-tumor tissues from 15 donors without cancer^37^ (see Methods). *RPL14* exhibited a universal expression across all cell types, notably in the malignant epithelial cells (**Figure 3**). Similarly, most HLA-I genes also showed broad expression in all cell types. By contrast, *SELE* exhibited specific expression in endothelial cells across both tumor and non-tumor tissues (**Figure 3C, Figure S4**). We also noted specific expression of some genes in cell types other than the malignant epithelial cells in NPC tumors (**Figure 3**). For example, *NOTCH3* was specifically expressed in cancer associated fibroblasts, while HLA-II genes were mainly expressed in various stromal cell types in the tumor microenvironment (TME; **Figure 3**). Additionally, by performing single-cell transcriptomic analyses of tumor samples from other epithelial cancers, including lung, gastric, and colorectal cancers (see Methods), we observed that gene associated with those cancers exhibited similar expression patterns observed in NPC, with some expressed predominantly in epithelial cells and some predominantly in stromal cells in the TME (**Figure S5**). These findings strongly indicate that genes conferring susceptibility to cancers are not necessarily confined to expression in malignant cells. Instead, these genes often exhibit predominant expression in diverse cell populations within the TME, suggesting a crucial role of genetic impact on stromal cells in cancer development.

**Figure 3.**
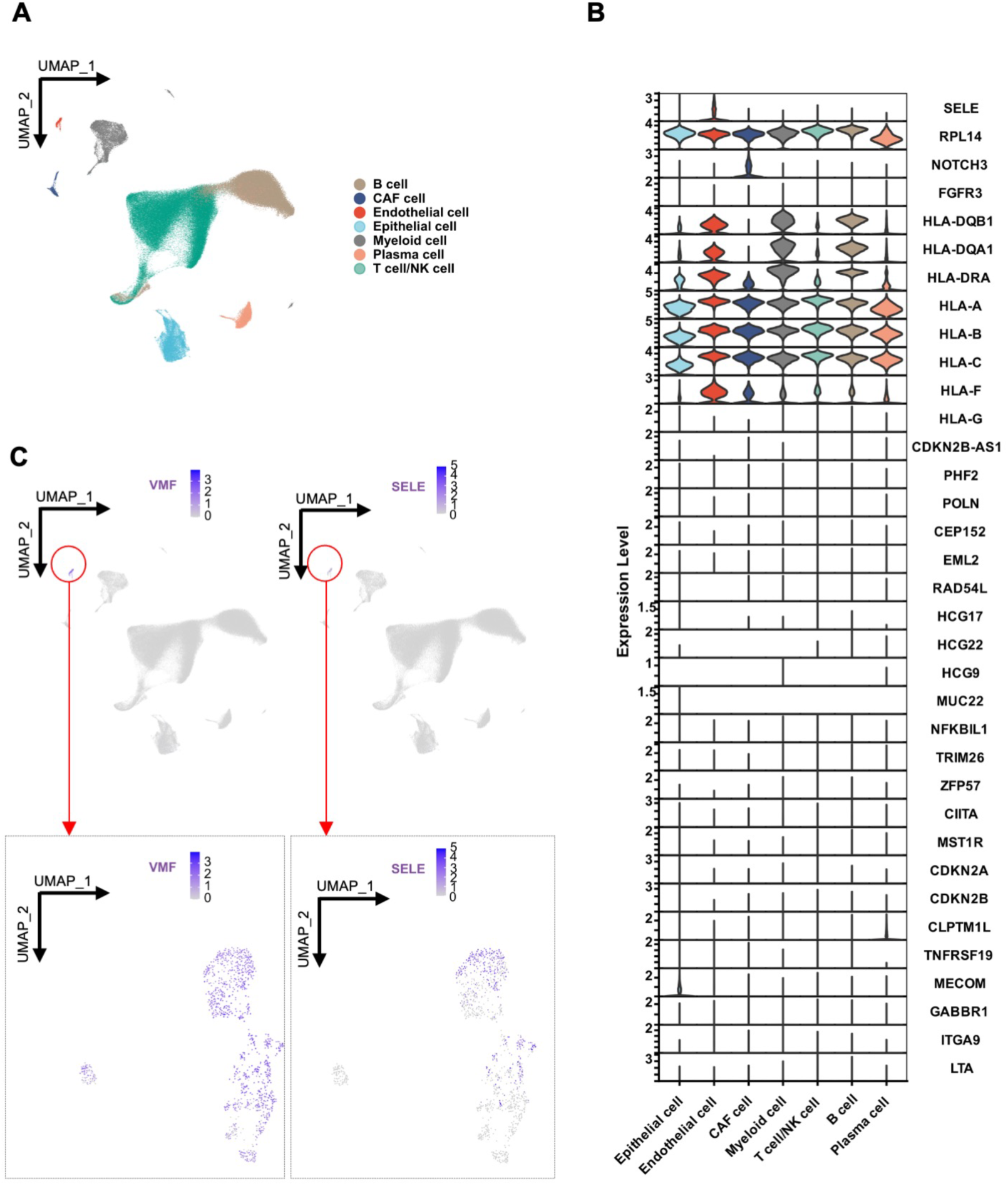
Expression patterns of novel and known NPC-associated genes across diverse cell types in NPC tumor tissues. Single-cell transcriptomic analyses of 223,593 cells derived from NPC tumor tissues in 56 patients. **A**. UMAP plot of 223,593 single cells grouped into seven major cell clusters. **B**. Violin plot illustrating normalized expression of NPC-associated genes across these major cell clusters. All epithelial cells captured in NPC tumor were malignant (see Methods). **C**. The expression of the marker gene (*VWF*) for endothelial cells and alongside the novel NPC-associated gene *SELE*; top panel: initial UMAP plot, bottom panel: re-normalized UMAP emphasizing cells highlighted by the red circles in the initial UMAP plot.

### rs2276868 regulates RPL14 expression through the NKRF transcription factor

Given the location of rs2276868 at the RPL14 promoter ^38^ (**Figure S6, S7A**) and the stable expression of *RPL14* in malignant epithelial cells (**Figure 3**), we aimed to investigate the potential regulatory role of rs2276868 in *RPL14* mRNA expression in epithelial cells. We first performed a single-cell-transcriptome-based eQTL analysis using 15,623 malignant epithelial cells from tumor tissues of 35 NPC patients. We observed that CC or CT genotype of rs2276868 was significantly associated with a higher expression of *RPL14* in malignant NPC cells compared to the TT genotype (**Figure 4A**). Subsequently, luciferase reporter assay demonstrated that the rs2276868-[T] construct had significantly lower luciferase activity compared to the rs2276868-[C] construct in human 293T cells (**Figure 4B**). These findings strongly suggest the influence of rs2276868 on *RPL14* transcription, which may involve modulating the binding affinity of certain transcription factor (TF).

**Figure 4.**
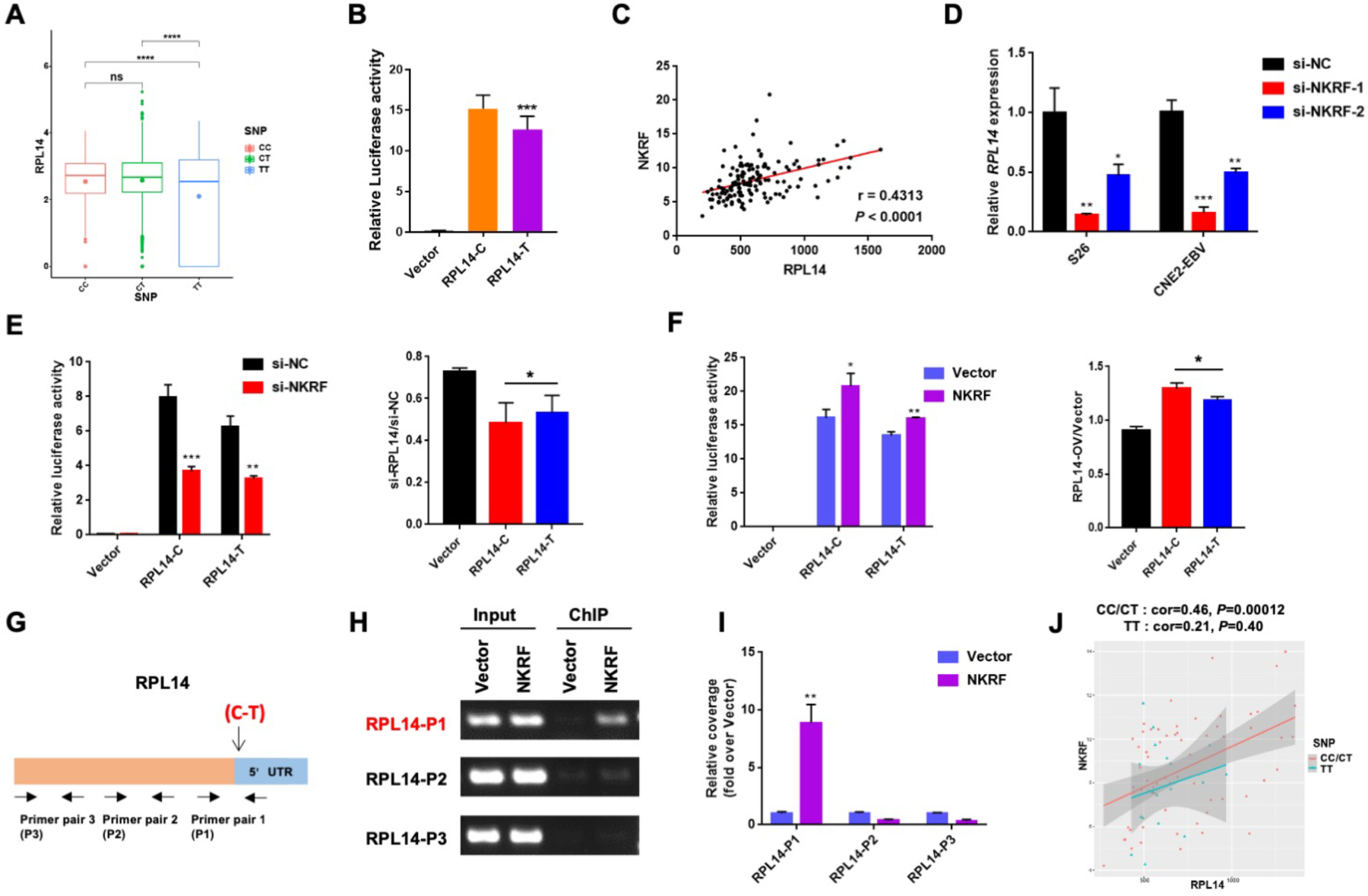
rs2276868 regulates the expression of RPL14. **A**. Single-cell transcriptome analysis showed the mRNA expression of RPL14 in 15,623 malignant epithelial cells from 35 NPC samples with different rs2276868 genotypes. **B**. Relative luciferase activity changes in 293T cells transfected with plasmids containing rs2276868 -[C], -[T], or control vectors. **C**. Pearson correlation analysis indicates the relationship between *RPL14* and *NKRF* expression (measured as transcript per million, TPM) in transcriptome data of NPC patients (n=87). **D**. RT-qPCR illustrates the mRNA expression of RPL14 in NPC cells transfected with NKRF siRNAs or control siRNA. **E-F**. Relative luciferase activity in 293T cells co-transfected with the rs2276868 -[C] or -[T] plasmids and NKRF siRNA (F) or NKRF overexpression vectors (G). Corresponding statistics are presented at the right. **G**. Schematic diagram indicates primer pairs used for PCR amplification of RPL14 fragments. **H-I**. ChIP assay in S26 cells transfected with Flag-NKRF and control vectors. ChIP PCR (H) and qPCR (I) showed RPL14 expression level in cells. P1-3 denote primer pairs targeting genomic regions shown in G. **J**. Pearson correlation analysis indicates the relationship between NKRF and RPL14 expression in single-cell transcriptome data are different for NPC patients with different genotypes. rs2276868 - [CC/CT] patients have a stronger correlation than rs2276868-[TT] patients. \**p* < 0.05, \*\**p* < 0.01, \*\*\**p* < 0.001.

To identify such potential TF, we conducted a comprehensive screening and identified 24 candidate TFs, which showed a positive correlation with the mRNA expression of *RPL14* in two independent bulk transcriptomic datasets of NPC tissues (n_Bei_=87 and n_Zhang_=113; **Figure 4B, Figure S7B**). Subsequent siRNA screening assays confirmed that five of the 24 TFs were capable of modulating *RPL14* expression (**Figure S7C**). Among them, *NKRF* knockdown significantly reduced *RPL14* transcription in NPC cells (**Figure 4B, Figure S7C-E**). Furthermore, knockdown or overexpression of *NKRF* exhibited a preferential effect on the transcription activity of the rs2276868-[C] construct over the rs2276868-[T] construct in luciferase reporter assays (**Figure 4E, F**). Consistently, single-cell transcriptome analysis revealed a stronger positive correlation between the expression of *NKRF* and *RPL14* in malignant NPC cells carrying the rs2276868-[C] compared to the rs2276868-[T] carriers (**Figure 4J**). Moreover, chromatin immunoprecipitation (ChIP) assay demonstrated that NKRF specifically binds to a refined genomic region encompassing rs2276868 (**Figure 4G-I, S7F**). Taken together, these data strongly suggest that rs2276868 influences *RPL14* expression through its interaction with *NKRF*, which preferentially binds to the promoter region housing rs2276868-[C] to foster enhanced transcription of *RPL14* in NPC tumors.

### RPL14 suppresses EBV infection and lytic cycle in NPC

To investigate the role of *RPL14* in NPC development, we first examined the relationship between *RPL14* expression and the transcriptional activity scores of various molecular pathways in NPC (see details in Methods). Through REACTOME and GO analysis of malignant epithelial cells in NPC scRNA-seq data (n_sample_=35 and n_cell_=15,623), we observed a significant association between *RPL14* and a pathway related to viral translation (GO:0019081: viral translation. P_sc_=1.63 × 10^-6^; **Figure 5A, Table S11**). Additionally, REACTOME and GO analysis of bulk RNA-seq data from NPC tissues (n_bulk_Bei_=93 and n_bulk_Zhang_=113) revealed significant associations between *RPL14* and multiple pathways related to viral infection. These pathways include SARS-COV-2 modulation of host translation machinery (P_bulk_Bei_=8.48 × 10^-16^, P_bulk_Zhang_=9.11 × 10^-14^) and influenza infection (P_bulk_Bei_=2.00 × 10^-14^, P_bulk_Zhang_=1.66 × 10^-13^; **Figure S8; Table S12, S13**). Considering the well-known involvement of EBV in NPC etiology^39^, we hypothesized a potential link between *RPL14* and EBV processes. Supportively, in EBV-positive malignant NPC cells, we observed a significant negative association (R= -0.27, P < 2.2×10^-16^) between *RPL14* expression and the EBV activity score, a metric developed by consolidating expression levels of multiple EBV genes (see Methods; **Figure 5B**). Furthermore, rs2276868-[T] carriers exhibited decreased *RPL14* expression but increased expression of EBV latent genes *LMP1* and *LMP2* as compared to rs2276868-[C] carriers in the same cell group (**Figure 5C**). These results corroborate the connection between *RPL14* and EBV-driven processes.

**Figure 5.**
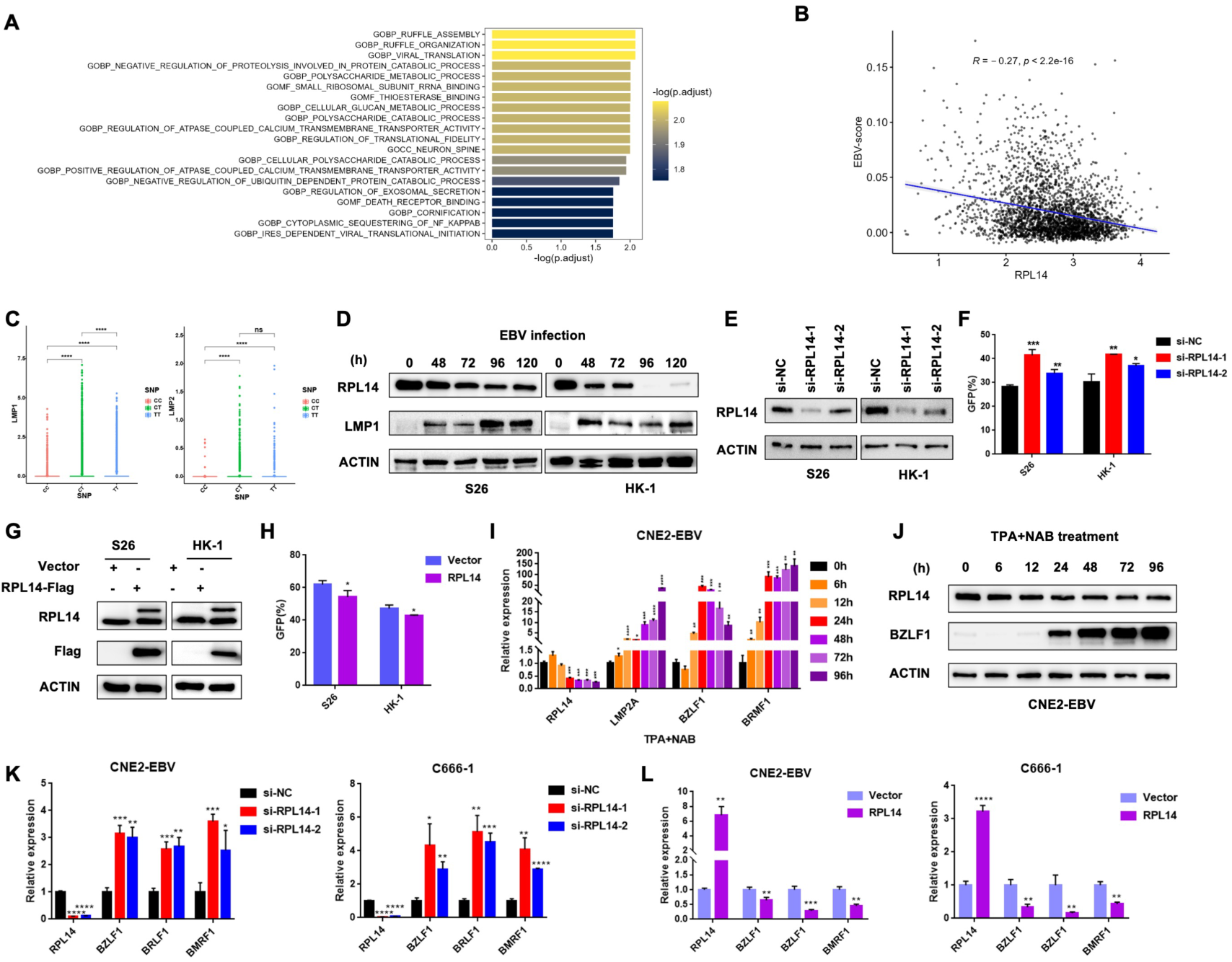
*RPL14* inhibits EBV infection and lytic cycle activation in NPC cells. **A.** The top 20 pathways significantly associated with RPL14 expression in malignant epithelial cells from NPC tumor. **B.** Correlation analysis between EBV-activity scores and *RPL14* expression within malignant epithelial cells. **C.** Single-cell transcriptome analysis showing *LMP1* and *LMP2* expression in NPC samples with either rs2276868 -[C] or -[T] genotypes. **D.** Western blot examination of RPL14 and LMP1 protein levels in NPC cell lines S26 and HK-1 cells infected with EBV at different timepoints. **E**. Western blot assessment of the knockdown efficiency of RPL14 siRNAs or control siRNA in S26 and HK-1 cells. **F.** Flow cytometry quantification of GFP intensity for the EBV infection efficiency in the NPC cells described in E. **G**. Western blot assay showing RPL14 protein expression in S26 and HK-1 cells infected with lentivirus stably expressing RPL14. ACTIN served as a loading control. **H**. Flow cytometry assessment of EBV infection efficiency in the cells described in G. **I-J**. Changes of gene expression in CNE2-EBV cells subjected to EBV lytic induction with TPA and NAB. RT-qPCR (I) and western blotting (J) were performed to detect the mRNA and protein expression, respectively, of RPL14 and EBV genes at different timepoints of EBV lytic induction. **K-L**. RT-qPCR analysis of EBV lytic gene expression in CNE2-EBV and C666-1 cells transfected with RPL14 siRNAs or control siRNA (K), or infected with lentivirus stably expressing RPL14 vector or control vector (L). \**p* < 0.05, \*\**p* < 0.01, \*\*\**p* < 0.001, \*\*\**p* < 0.0001.

To further explore the interaction between *RPL14* and EBV, we monitored the protein expression of RPL14 in NPC cells post EBV infection over different timepoints. We observed a significant downtrend in RPL14 expression concurrent with EBV infection (**Figure 5D**). By contrast, RPL14 knockdown resulted in a significant rise in EBV infection in NPC cells, while RPL14 overexpression led to a marked reduction in EBV infection (**Figure 5E-H**). We also noticed a decline in RPL14 expression when EBV lytic cycle was induced using TPA and NaB in NPC cells (**Figure 5I, J; Figure S9**). Furthermore, RPL14 knockdown remarkably augmented the expression of EBV lytic genes in NPC cells, while RPL14 overexpression significantly reduced their expression (**Figure 5K, L**). These observations strongly suggest that RPL14 acts as a suppressor of EBV infection and lytic cycle in NPC.

### RPL14 impedes tumorigenesis in NPC cells

We next explored the function of *RPL14* in NPC tumorigenesis. We first manipulated *RPL14* expression in an *in vitro* NPC cell model (**Figure 6A; S10A**). Cell growth curves and colony formation assays revealed that the stable overexpression of *RPL14* significantly inhibited NPC cell proliferation. Conversely, stable knockdown of *RPL14* had an opposite effect, promoting proliferation (**Figure 6B-C, Figure S10B-C**). Transwell assays revealed that *RPL14* overexpression suppressed NPC cell migration, while its knockdown enhanced the migration (**Figure 6D; Figure S10D**). These observations strongly suggest that *RPL14* inhibits the tumorigenesis of NPC cells.

**Figure 6.**
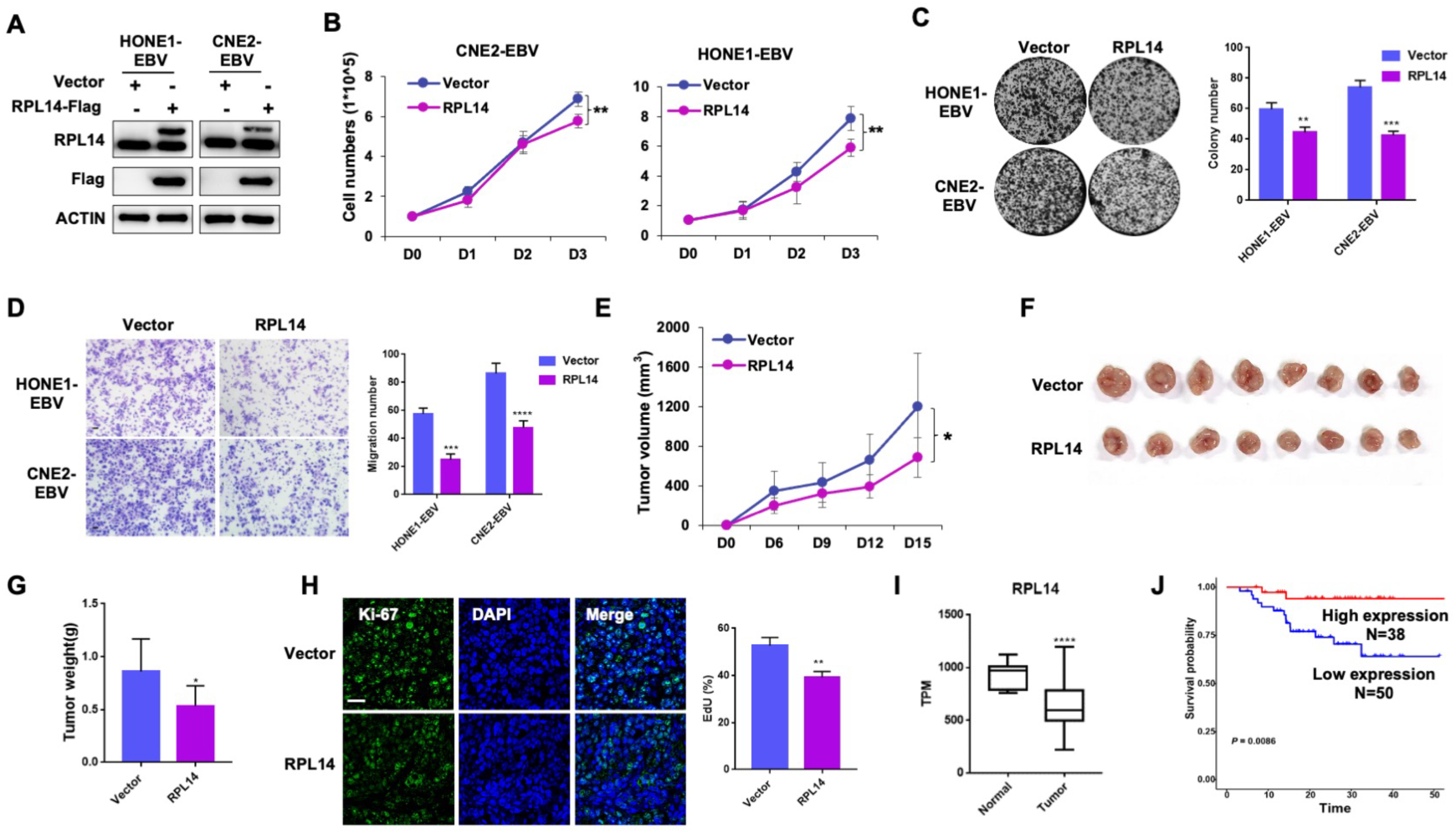
Tumor suppressive function of *RPL14* in NPC. **A**. Western blot analysis showing RPL14 protein levels in NPC cells infected with RPL14 overexpressing lentivirus. ACTIN serves as the loading control. **B**. Cell growth curves of the cells described in A. **C**. Colony formation assay for the cells described in A. Corresponding statistical analysis is shown to the right. **D**. Transwell migration assay evaluating the migration ability of the cells described in A. The statistical analysis is presented on the right. **E-H**. Tumor growth evaluation in a nude mice model with subcutaneous injection of NPC cells described in A. Tumor volumes were measured every three days. Visual presentation of tumor post-sacrifice (F) and weight (G) were presented. IF detection of Ki-67 expression (H) in the tumors described in F, and the corresponding statistical analysis is shown on the right. **I**. Transcriptomic analysis showcasing mRNA levels of RPL14 in NPC (n=87) versus control samples (n=10). **J**. Kaplan-Meier survival analysis linking RPL14 expression to survival time in NPC patients using bulk RNA-seq data (n=88). \**p* < 0.05, \*\**p* < 0.01, \*\*\**p* < 0.001, \*\*\*\**p* < 0.0001.

To corroborate the tumor-suppressive function of *RPL14 in vivo*, we established a mouse model with subcutaneous injection of NPC cells stably expressing *RPL14*. We observed a significant inhibition on tumorigenesis as reflected by the diminished tumor volume and weight in mice with *RPL14* overexpression compared to the control group (**Figure 6E-G**). Supportively, immunofluorescence staining of Ki-67, a proliferation marker, revealed a substantial decrease in Ki-67 positive cells in the *RPL14*-overexpressing tumors compared to the control tumors (**Figure 6H**). Furthermore, we observed a significant lower expression of *RPL14* in NPC tissues compared to nasopharyngeal tissues from individuals with rhinitis (**Figure 6I**). Kaplan-Meier survival analysis revealed that low expression of *RPL14* was significantly correlated with a poor prognosis of NPC patient (**Figure 6I**). These findings strongly suggest *RPL14* as a tumor suppressor in NPC.

### SELE-S149R promotes NPC development through gain of function in endothelial cells

*SELE* encodes E-selectin, a selectin cell adhesion molecule specifically expressed on the surface of endothelial cells (**Figure 3C**). E-selectin plays a pivotal role in attracting cancer cells by binding to ligands like glycoproteins^40^. Structural modeling of E-selectin paired with glycomimetic antagonist ligand revealed that the Ser149 residue of SELE, implicated in the rs5361 variant, is in close proximity to its ligand, and the S149R substitution (**Figure 2C**) leads to an extension of E-selectin’s side chain, potentially altering its interaction with glycoproteins (**Figure 7A**). This aligns with a previous finding that *SELE-*S149R introduces a gain-of-function effect on the recruiting capability of E-Selectin^41^.

**Figure 7.**
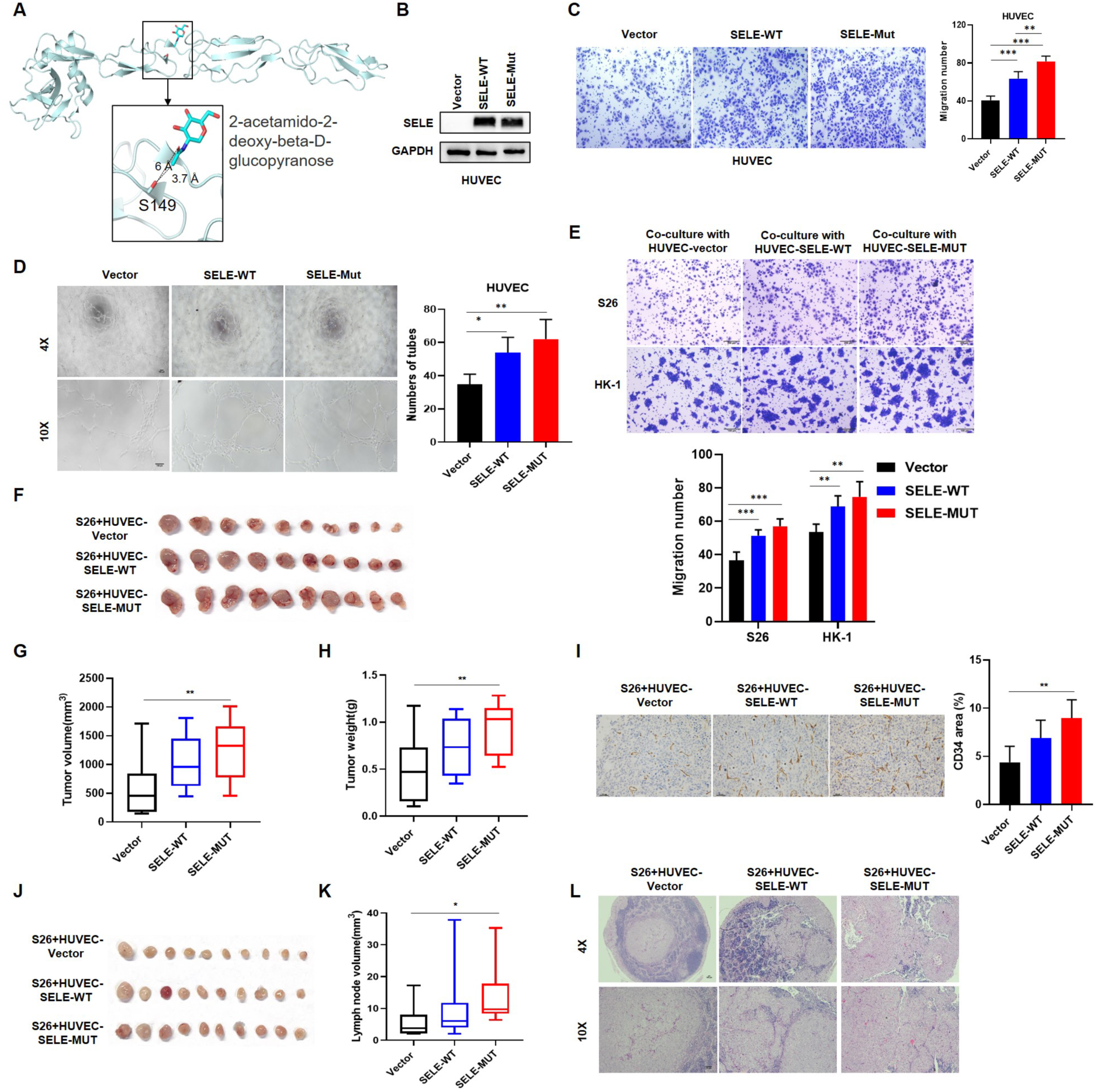
SELE-S149R mutation in endothelial cells promotes the tumorigenesis and metastasis of NPC cells. **A.** Structure prediction of E-selection paired with glycomimetic antagonist ligand 2-acetamido-2-deoxy-beta-D-glucopyranose (PDB code 4C16). The spatial proximity between Ser149 and the ligand is highlighted. Both the Ser149 side chain and the ligand are shown as sticks, with the distances between the OG atom of Ser149 and the C8 or O7 atoms of the ligand specified. **B.** Western blot examination of SELE protein level in HUVEC cells infected with lentivirus overexpressing SELE-WT, S149R mutant or control vectors. **C.** Transwell migration assay evaluating the migration ability of the cells described in B, with statistical analysis presented at the bottom. Scale bar, 100 μm. **D.** Tube formation assay with cells described in B. The statistical analysis is presented on the right. Scale bar, 100 μm. **E.** Transwell migration assay assessing the migration ability of the NPC cell lines (S26 and HK-1) co-cultured with HUVEC cells from B (S26/HK-1: HUVEC = 10:1), accompanied by statistical analysis presented at the bottom. Scale bar, 200 μm. **F-H.** Tumor growth evaluation in xenograft model with subcutaneous injection of S26 cells described in E. Tumor volumes. (G) are measured and visual presentation of tumor post-sacrifice (F) and weight (H) are presented. **I.** IHC staining of CD34 in tumors presented in F. The statistical analysis is presented on the right. Scale bar, 100 μm. **J-L.** Tumor lymphatic metastasis of S26 cells described in E. Lymph nodes are visualized (I) and measured (J), with HE staining conducted to assess metastasis in these lymph nodes (K). Scale bar, 100 μm. \**p* < 0.05, \*\**p* < 0.01, \*\*\**p* < 0.001.

To probe the functional implications of the *SELE-*S149R mutation in the context of NPC, we developed a model using human umbilical vein endothelial cells (HUVECs) stably overexpressing either the wild-type SELE (*SELE*-Ser149, SELE-WT) or its S149R mutant variant (SELE-MUT, **Figure 7B**). Transwell assays revealed that overexpression of SELE, especially the SELE-MUT, significantly boosted the migration ability of HUVEC cells (**Figure 7C**). Tube formation assay further demonstrated that SELE, particularly the mutant form, markedly enhanced the endothelial cell’s ability to form capillary-like structures, (**Figure 7D**), indicating a gain-of-function effect of the S149R mutation in promoting angiogenesis. We next employed a co-culture model involving HUVECs (expressing either SELE-WT or MUT) and NPC cell lines (HK-1 and S26). Strikingly, HUVECs overexpressing SELE, especially the SELE-MUT variant, substantially increased the migration of NPC cells (**Figure 7E**). This was further substantiated by *in vivo* xenograft assays, which showed increased tumorigenesis in NPC cells co-cultured with HUVEC-SELE-MUT, evident from the augmented tumor volume and weight (**Figure 7F-H**). Moreover, IHC staining of CD34, a marker of vessels, revealed that SELE-MUT overexpression significantly promote the angiogenesis *in vivo* (**Figure 7I**). Furthermore, lymph node metastasis assays reinforced these findings, demonstrating that co-culture with HUVEC-SELE-MUT cells substantially promoted the metastasis of NPC cells (**Figure 7J-L**). These results strongly suggest that *SELE-*S149R mutation in endothelial cells plays a crucial role in escalating both tumorigenesis and metastasis in NPC cells.

### Genetic contributions of common and rare variants to NPC susceptibility

We next undertook a comprehensive analysis to quantify the overall heritability of NPC explained by SNVs derived from WES, which was further broken down based on the MAF and LD score classifications of these SNVs, using GREML-LDMS model (see Methods). We observed that a total of 15.2% of NPC susceptibility (with a standard error of 3.6%) could be explained by the genetic effects of all WES-derived SNVs, including 4.4% and 10.8% attributed to variants within the HLA and non-HLA regions, respectively. Notably, rare variants (MAF < 0.01) in the non-HLA regions explained a substantial 82.4% of the WES SNV-heritability (**Figure 8A**). Furthermore, variants with the lowest LD scores, meaning relative independence, contributed the largest proportion (30.6%) to the WES SNV-heritability (**Figure 8A**).

**Figure 8.**
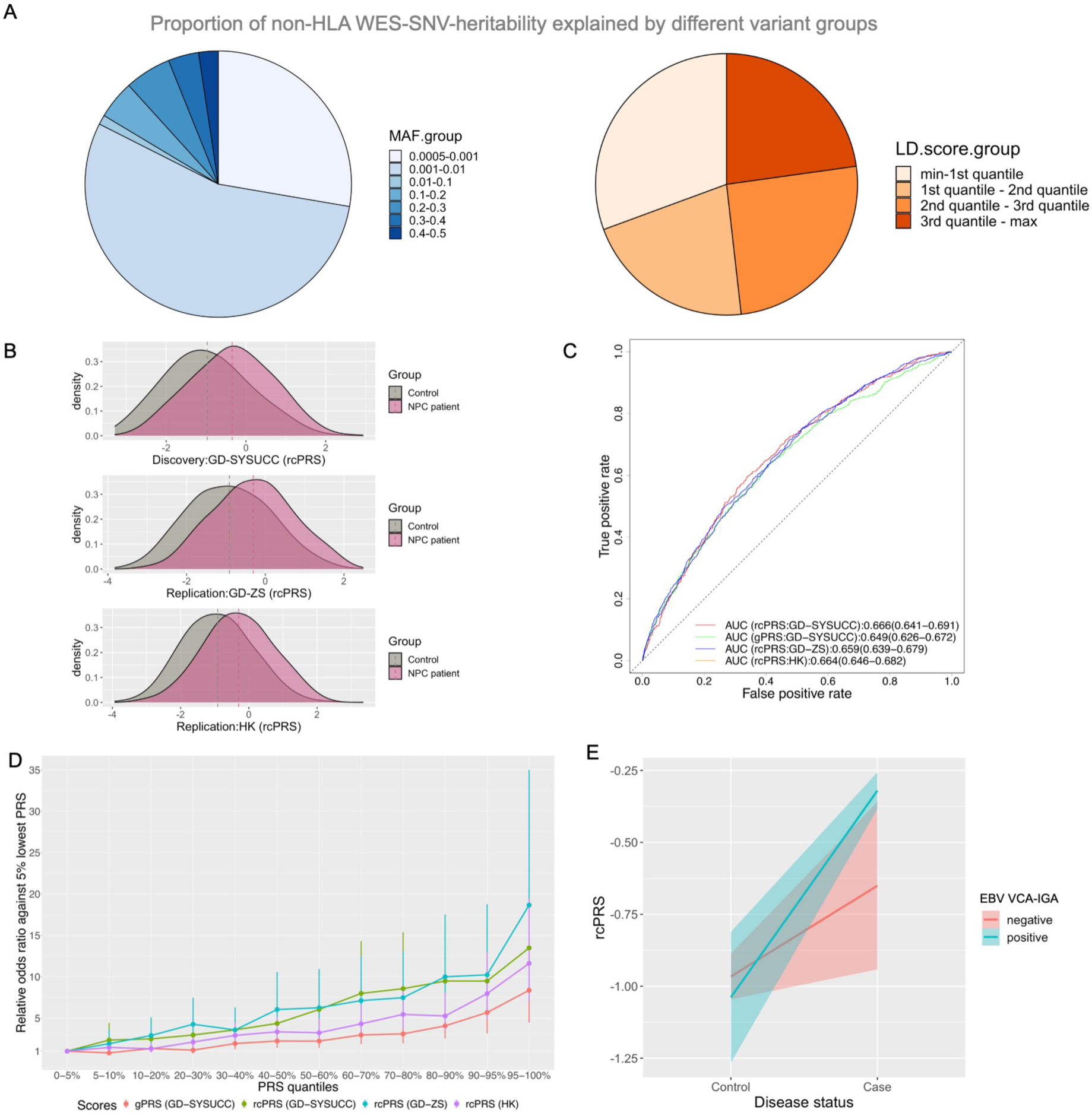
Contribution of common and rare variants to NPC susceptibility. **A**. Fractional representation of NPC heritability attributable to non-HLA WES-SNVs, categorized by minor allele frequency (MAF) and linkage disequilibrium (LD). For each variant, MAF was calculated using the discovery samples, and LD score represented the aggregated R^2^ with adjacent variants spanning a 200kb window. **B**. Density plots illustrating the PRS incorporating the novel loci and previously known GWAS loci (refined composite PRS, rcPRS) for cases and controls in both the discovery and replication cohorts. **C**. Receiver operating characteristic curves comparing the rcPRS and previously reported GWAS PRS (gPRS) for NPC across different cohorts. **D**. Relative odds ratio comparing the rcPRS or gPRS bins and the 5% lowest quantile group in different cohorts. **E**. Correlation of rcPRS with disease status in individuals categorized by their EBV VCA-IgA status.

Through a comparative analysis using a joint model that considers multiple genetic risk factors simultaneously (see Methods), we further revealed that the minor allele G of the rare variant *SELE*-rs5361 conferred a substantial disease risk (OR=2.20∼2.22, **Figure S11**) in the two Guangdong cohorts (GD-SYSUCC and GD-ZS). This risk level is comparable to that associated with the highest 20% of the population’s polygenic risk score (PRS) derived from HLA variants (OR=2.14∼2.30, **Figure S11**). Intriguingly, the risk also overshadowed that conferred by the minor allele T of the common variant *RPL14***-**rs2276868 (OR = 1.22∼1.45) or the highest 20% of the population’s PRS for NPC from non-HLA GWAS variants (OR = 1.31-1.80, **Figure S11**). Additionally, by assessing the phenotypic variance ascribed to each locus^42^, we also revealed that HLA loci are predominantly responsible for the NPC phenotypic variance, while the *RPL14* common variant significantly explain a greater portion of disease variance than the *SELE* rare variant (**Figure S11**).

### Polygenic risk prediction improved by novel loci and mediated by EBV infection

To assess the efficacy of the novel loci in predicting NPC risk, we developed a refined composite polygenic risk score (rcPRS) that simultaneously considers NPC-associated SNVs and the leading SNVs in NPC-associated genes and pathways identified in our study and previous GWASs (see Methods; **Table S15**). Notably, the rsPRS demonstrated a consistent right-skewed distribution for NPC cases compared to controls across all discovery and replication datasets (**Figure 8B**), suggesting a higher predictive value of rsPRS in identifying individuals at risk. Furthermore, the rcPRS achieved an area under curve (AUC) for risk prediction ranging between 0.659 and 0.666 in the GD-SYSUCC discovery and replication samples, outperforming the published GWAS-based PRS in the GD-SYSUCC sample (gPRS; AUC = 0.649; **Figure 8C; see Methods**)^10^. This improvement in predictive accuracy is largely attributable to the inclusion of two novel common variants, rs1050462 in *HLA-B* and rs2276868 in *RPL14*, as evidenced by our leave-one-out analysis (**Table S16**). Further stratification of individuals based on their NPC PRSs, either rcPRSs or gPRSs, revealed a more pronounced rising trend in the relative disease risk with the escalating rcPRS compared to the gPRS (**Figure 8D**). Individuals in the highest percentile rage (95-100%) of the rcPRS exhibited approximately double the relative risk (OR_rcPRS_discov_=15.1) than those with the gPRS (OR_gPRS_discov_=8.4; **Figure 8D**). These findings suggest that incorporating novel loci into the rcPRS not only improves its accuracy in distinguishing disease risk but also enhances its capability to stratify individuals based on their likelihood to develop NPC.

Noteworthily, in a subset of 1,018 cases and 774 controls from the GD-SYSUCC cohort that had data on serological antibody against the EBV-encoded virus capsule antigen (EBV VCA-IgA), we observed a stronger association between rcPRS and NPC susceptibility in individuals tested seropositive for EBV VCA-IgA compared to those tested seronegative **(Figure 8E)**. Interaction analysis statistically further confirmed that the association between the rcPRS and NPC disease status was significantly mediated by EBV status (P_interaction_=0.03). These findings suggest that the polygenic risk of NPC is mediated by serum EBV activity, and that rcPRS may have a greater utility in predicting NPC risk among individuals with serological evidence of EBV infection.

## Discussion

Here we conducted the largest whole-exome sequencing association study on NPC to date and unveiled three novel variants linked to NPC susceptibility, including the common variant rs2276868 in the promoter (5’-UTR) region of *RPL14*, the rare coding variant rs5361 in *SELE*, and another common variant rs1050462 in HLA-B locus^8,11^. Notably, the identification of the *RPL14* locus underscores the significant role of familial history in NPC. This mirrors patterns observed in other complex diseases^42^, suggesting both genetic or environmental factors related to family history might amplify the genetic impact of individual variants on NPC risk. A possible explanation for this locus remaining undetected earlier could be its pronounced genetic risk in familial cases (OR_FHNPC_ =1.575) compared to sporadic cases (OR_sporadic_NPC_ = 1.200), coupled with the vast sample size of our study. Additionally, the successful replication of associations with *SELE* and other two sentinel pathway genes underlines an efficient approach to identify and decompose disease associations at the pathway level.

Our study sheds light on the functional roles of the novel genes, offering a better understanding of the pathogenic mechanisms underlying NPC. We uncovered that the variant rs2276868 modulates *RPL14* expression through the transcription factor *NKRF*, which is an inhibitor to the NF-κB pathway involved in NPC development^43,44^. Single-cell transcriptomic analyses revealed an inverse association between *RPL14* expression and EBV activity in malignant NPC cells, which is further corroborated by the suppressive effect of RPL14 on EBV infection and lytic cycle in NPC cells. Given the involvement of RPL22^45^ and RPL4^46^, both of which belong to the same RPL family as RPL14^47^, in EBV infection, these findings highlight the crucial role of RPL family members in EBV life cycle. Furthermore, our study revealed that *RPL14* inhibits NPC tumorigenesis, consistent with its tumor-suppressive function reported in an *in vitro* NPC study^48^ and in other cancer types^49^. Reinforcing this, a high *RPL14* expression is associated with a favorable survival of NPC. Taken together, our findings suggest that the polymorphism at rs2276868 predisposes individuals to varied *RPL14* expression, which in turn influences EBV-related tumorigenesis in NPC.

Besides *RPL14* in our study, multiple NPC-associated genes such as *HLA-I, -II*, and *CIITA* have been implicated in modulating EBV life cycle ^29,30^. This reinforces that genetic variations affect EBV to mediate varied susceptibility to NPC. Significantly, our findings reveals that the effectiveness of the NPC polygenic risk score is influenced by the serological status of EBV activation. Given that both the EBV DNA load and antibodies against EBV are established serological markers for early NPC diagnosis^60,61^, incorporating the rcPRS and EBV infection markers could significantly enhance personalized risk assessments and lead to the development of more effective screening strategies for NPC.

Our study has highlighted *SELE* as a novel NPC-associated gene. This association is likely attributed to the rare germline mutation rs5361 in *SELE*, which encodes an adhesion protein E-Selectin, known for its interactions with various cells, including leukocytes and malignant epithelial cells^36,50^. A previous study has demonstrated that the deleterious S149R encoded by rs5361 enhances the functionality of E-Selectin in endothelial cells, promoting their rolling and adhesion capabilities^37^. Noteworthily, our study extends these findings by demonstrating that *SELE* expression is confined to endothelial cells in tumor tissues. Our structural modelling and functional experiments reveal that S149R mutation extends the SELE’s side chain on the surface of endothelial cells, thereby augmenting their migration and angiogenesis abilities. Furthermore, we observed that endothelial cells expressing SELE, particularly those with the S149R mutation, significantly render stronger migration and tumorigenesis to malignant NPC cells. This supports the crucial role of E-selectin in interacting with cancer cells and modulating metastasis across various cancers^36,51–53^. Therefore, these findings suggest that the *SELE-* S149R may enhance the adhesion function of E-Selectin in endothelial cells, reshaping the key TME component to favor the progression of cancer cells^53^, consequently altering individual susceptibility to NPC.

We further delved deeper into the cellular expression of various NPC-associated genes, revealing versatile patterns. For example, certain genes, like *RPL14*, are expressed in most cell types, including epithelial cells that give rise to malignant NPC cells. In contrast, genes like *HLA-II* are found mainly in diverse stromal cells, which typically lack malignant potential in NPC. Such patterns are also observed in other cancers. For instance, the well-known susceptibility-associated genes *NOTCH4* in lung cancer and *SMAD1* in colorectal cancer^54,55^ are exclusively expressed in endothelial cells (not typically developing into malignancy) in tumors. These findings highlight the multifaceted cellular implications of germline variations in cancer susceptibility, resonating with a growing literature suggesting the engagement of various cell types in the development of complex disease^56–58^. Given that the validation of the functional relevance of genetic susceptibility genes often predominantly emphasizes the cells of cancer origin (e.g., epithelial cells in NPC), our findings call attention to the importance of exploring the roles of cancer risk genes across diverse TME cell types in cancer development (such as endothelial cells)^54,55^.

In summary, our study fills the missing heritability of NPC and enhances our understanding of the genetic architecture of NPC by unveiling novel genetic associations from variants of both common and rare prevalence. Furthermore, our study elucidates diverse pathogenic mechanisms, including the EBV-mediated tumorigenesis in malignant epithelial cells through *RPL14* and the adhesion component in endothelial cells via *SELE*. These findings highlight the substantial impacts of genetic variants on EBV life cycle and various cell types within the TME, which jointly contribute to NPC development. Moreover, our innovative rcPRS model, incorporating these novel discoveries, reflects the modifier effect of EBV status on polygenic risk and provides promising implications for NPC screening. Additionally, we acknowledge limitations, including sample size constraints, especially for the FHNPC group and rarer variants, and the need for further studies to delve into the mechanistic links between these genetic variants and NPC pathogenesis. Future work should explore the potential clinical benefits of these findings in NPC management.

## Data Availability

Key data were deposited in the Research Data Deposit public platform (RDD; http://www.researchdata.org.cn).

## Acknowledgement

We acknowledge supports from the National Natural Science Foundation of China (NSFC; 82261160657, 81971270), the National Key R&D Program of China (NKRDPC; 2022YFC3400901 and 2021ZD0202000), the Sci-Tech Project Foundation of Guangzhou City (201707020039), the Guangdong Innovative and Entrepreneurial Research Team Program (2016ZT06S638), the Chang Jiang Scholars Program (J.X.B.), Hong Kong Research Grant Council (RGC) Theme-based Research Scheme Funds (T12-703/22-R and T12-703/23-N), National Medical Research Council Singapore Clinician Scientist Award (NMRC/CSAINV20nov-0021), the Duke-NUS Oncology Academic Program Goh Foundation Proton Research Programme (M.L.C), NCCS Cancer Fund (M.L.C), the Kua Hong Pak Head and Neck Cancer Research Programme (M.L.C), the Research Grants Council Area of Excellence (AoE) Hong Kong NPC Research (AoE/M-06/08), and the Hong Kong Cancer Fund (M.L.L.). We thank all participants in the study, staffs at the biobank of SYSUCC for sample preparation, staffs at the Tissue Bank for providing NPC blood samples and the patient demographics of the HK cohort, Tan Kah Min’s support for technical support and sample processing in SG cohort, and staffs at the High-Throughput Analysis Platform (HTAP) of SYSUCC for data generation and processing.

## Author contributions

Designed the study: J.X.B, Y.X.Z; procured financial support: J.X.B, M.L.C, M.L.L, M.J; sample recruitment, preparation, and data collection: M.J, F.G.L, C.C.K, Z.L, M.L.L, J.M.K, M.L.C, E.H.O, Y.M.G, J.R.C, S.H, S.Q.L, X.X.C, X.Y, B.W, Y.H.Z, A.Y.X, P.P.W, Q.Y.C, L.Q.T, W.H.J, H.Q.M; analyzed and interpreted data: Y.Z, J.X.B, C.L.L, G.W.L, X.B, Y.L, S.H, Z.H.R, Y.L.C, C.C.K, J.J.L, M.L.C, J.M.K, E.H.O, J.D.M, D.X.L; performed functional experiments: C.L.L, J.X.J, W.X.Y, Y.Q.Z; wrote the paper (original draft): Y.Z, C.L.L, J.X.B; all authors approved the final report.

## Materials and Methods

### Ethics

The study was approved by the Sun Yat-sen University Cancer Center (SYSUCC) Ethics Committee (reference no. SL-B2021-032-03), the SingHealth Institutional Review Board (IRB protocol no. 2019/2177), and the Institutional Review Board (IRB) of the University of Hong Kong. Informed consent was obtained from all participants.

### Patient recruitment and sample preparation

#### Discovery stage

A total of 3,039 NPC patients of Chinese Han ancestry was recruited from Guangzhou in Guangdong province of China (GD-SYSUCC; n= 2,196) by SYSUCC and from Singapore (SG; n=843) by the National Cancer Center (Singapore, collected from 1987-2018). A total of 2,692 healthy individuals of Chinese Han ancestry was recruited as controls from local community in Guangdong (GD-SYSUCC; n=1,355) and from Singapore (SG; n =1,337) by Genome Institute of Singapore (Singapore, details have been described previously^63^). All NPC cases were histologically diagnosed. All healthy controls declared free of cancer through questionnaires. After stringent quality control, the discovery dataset consisted of 2,694 NPC cases (2,125 from GD-SYSUCC and 569 from SG) and 2,328 healthy controls (1,068 from GD-SYSUCC and 1,260 from SG). Serum immunoglobulin A (IgA) antibodies to Epstein-Barr virus (EBV) capsid antigen (IgA-VCA) were available for 1,646 NPC patients and 918 controls in GD-SYSUCC subset. Individuals with IgA-VCA >=1:20 was categorized as EBV positive.

#### Replication stage

The replication dataset included two cohorts: a Zhongshan cohort (GD-ZS) consisting of 1,941 cases and 2,265 controls recruited from the Zhongshan city in the Guangdong province by Zhongshan City People’s Hospital, and a Hong Kong cohort (HK) consisting of 2,334 cases and 2,507 controls recruited from Hong Kong by the University of Hong Kong. Specifically for the HK cohort, NPC cases and controls were collected by the Tissue Bank established under the Area of Excellence (AoE) program from five Hong Kong public hospitals including Queen Mary Hospital (QMH), Queen Elizabeth Hospital (QEH), Tuen Mun Hospital (TMH), Pamela Youde Nethersole Eastern Hospital (PYNEH), and Princess Margaret Hospital (PMH) from 2010 to 2017, details have been described previously^11^. NPC cases were confirmed by histopathology and enrolled with routine staging procedures according to the American Joint Committee on Cancer (AJCC) TNM system, physical examination, and imaging tests. The control population was recruited from the Red Cross and hospital cancer-free individuals from QMH, QEH, TMH, PYNEH, and PMH. We considered GD-ZS as a Guangdong indigenous cohort that shared regional demographic features with the discovery GD-SYSUCC cohort collected from the same province, whereas the HK cohort shared ancestral genetics with other cohorts but was relatively demographically different.

#### Bulk RNA-Seq samples

Tumor biopsy samples from 93 NPC patients were collected for bulk RNA-seq (labelled as “the Bei-lab collection”), 87 of which had prognosis data available. Additional bulk RNA-seq data of tumor tissues from 113 independent NPC patients with prognosis data available were collected from a published study (labelled as “the Zhang-lab collection”)^64^. We further collected nasopharynx samples from 10 cancer-free individuals with inflammations in nasopharynx at SYSUCC, which were used as controls. Biopsies were collected before any treatment and immersed in RNAlater.

#### NPC samples for Single-cell RNA-seq

Tumor biopsies of 10 NPC patients were collected for single cell RNA sequencing (scRNA-seq) in our previous study^32^. Among them, eight had tumor epithelial cells successfully captured. By extracting data from four additional studies^33,65–67^, we further collected scRNA-seq data of 46 NPC tumor samples, 27 of which captured tumor epithelial cells^33,65–67^. In total, scRNA-seq datasets consisting of 56 samples (of which 35 have epithelial cells captured) were used in downstream analyses.

### Data generation, quality control and annotation

#### Whole exome sequencing and variant calling

Whole blood DNA sample was extracted from all participants in the discovery dataset for whole exome sequencing. Library was constructed using one of the three products: Agilent SureSelect Human All Exon V6+UTR kit, Agilent SureSelect Human All Exon V5+UTR kit, and Roche SeqCap EZ Exome + UTR Target Enrichment Kit. This was followed by sequencing using a paired-end 2 x 150 bp protocol on an Illumina HiSeq instrument. A custom optimized version of Sentieon^68^ was applied to preprocess sequencings reads, align reads to human genome using GRCh37 (hg19) assembly as reference, and conduct single nucleotide variant (SNV) calling using algorithms following the GATK best-practice pipeline^69,70^. After GATK’s VQSR quality control, hard filtering was applied to remove the variants with 1) Read depth (DP) <10; 2) Allelic Balance (AB) > 0.8 or < 0.2 for heterozygous genotype; 3) Genotype quality (GQ) < 30. For rare variants (minor allele frequency < 0.01), we applied additional stringent quality control based on per-read mutation rate, allelic balance, and strand bias (defined as proportion of allele called by reads from one strand over that by any of the two strands). We used 1000 genome phase 1 high confident SNP set as golden set to establish parameter threshold range where high liability variants can be called. This led to the following filtering for rare variants: 1) refining alignment by removing any read with a mismatch distance against the reference larger than 8 bp (any indel event counts for 1 bp) and re-apply hard filtering as described above; 2) setting variant-sample elements with strand bias > 0.95 or < 0.05 for heterozygous genotype, or >0.97 or <0.03 for homozygous genotype to missing; 3) setting variant-sample elements with allelic Balance (AB) > 0.75 or < 0.22 for heterozygous genotype, or with allelic Balance (AB) < 0.95 and > 0.04 for homozygous genotype to missing.

#### Additional quality control for association test

For both common and rare variants, we further removed variants that are 1) with a missing rate >= 0.02; 2) with a Hardy-Weinberg equilibrium P value <= 1 x 10^-6^ in healthy controls; 3) with a significant differential missing rate between case vs controls (Bonferroni corrected); 4) variants in super duplicates or low complexity regions. Samples were removed if they are 1) with a missing rate > 0.05; 2) with autosomal heterozygosity deviation outside 3 standard deviations of the mean; 3) genetic outliers through eye check or with any the of top four principal component value located outside 6 standard deviations of the mean. These filtering steps were performed using PLINK^71^. We also removed one of each related pair (remained sample t<0.05) using GCTA^72^.

#### HLA genotyping

High resolution WES-based HLA typing was performed on the bam files using HLAScan (v2.1.3) with default parameters and implemented HLA database^73^. PCR amplification HLA typing was also performed on 135 WES samples to cross-validate the WES-based typing procedures. The concordance of the HLA genotypes was high (> 94%) at four-digit resolution level for most HLA genes, except for HLA-DQA1 and -DPB1 with a relatively lower concordance being observed (> 80%). The HLA amino acid (AA) polymorphism was obtained using HLA AA alignments in IPD-IMGT database^74^. The genotypes of HLA SNPs were inferred by the SNP2HLA software^75^.

#### Capture sequencing for targeted genomic regions

DNA library preparation was performed as described in the NadPrep DNA Library Preparation Kit (#1002222; Nanodigmbio, Nanjing, China) for the MGI sequencing platform on the MGISP-960 automated workstation (MGI Tech Co., Ltd., Shenzhen, China), in accordance with the manufacturers’ instructions. Hybridization capture-based target enrichment was performed as described in the NadPrep Hybrid Capture Reagents kit (#1005101; Nanodigmbio) on the MGISP-100 automated workstation (MGI Tech Co., Ltd., Shenzhen, China), in accordance with the manufacturers’ instructions. The library distribution was analyzed by Qsep100 (BiOptic Inc, New Taipei City, China) and quantified by Qubit dsDNA HS Assay Kit (Invitrogen, Termo Fisher Scientifc Inc, Waltham, USA). Final libraries were sequenced on MGI DNBSEQ-T7 (MGI Tech Co., Ltd., Shenzhen, China) using a pair-end of 150 bp.

#### Whole transcriptomic sequencing

For bulk RNA-seq, total RNA was extracted from tissue or cell line using Rneasy Mini Kit (Qiagen), with ribosomal RNAs removed by Ribo-Zero Magnetic kit (Illumina). TruSeq RNA Library Prep Kit (Illumina) was used to construct the library, followed by RNA sequencing. RNA-seq was conducted with a pair-end of 150 bp protocol on an Illumina Hiseq X sequencer. Bowtie was used to map post-Qced reads to human reference genome (hg19) and Htseq was used to quantify reads counts^76,77^. Per-gene expression level was normalized using transcripts per million (TPM). scRNA-seq was performed following the manufacturers’ instructions (10x Genomics single-cell 5’ sequencing and 3’ sequencing kits). Details were described in previous studies^32,33,65–67^.

### Variant annotation

ANNOVAR was used to annotate SNV with following categories^78^:

1) SNPs in dbSNP150 database.
2) Variants in population genetics database: 1000 genome phase 3, exome and genome subset of the Genome Aggregation Database (EXAC, gnomAD).
3) within genes in NCBI RefSeq Gene database.
4) within genes of pathways in curated canonical pathways, GO Ontology items and oncogenic signature gene sets as downloaded from Molecular Signatures Database v6.2^79^.
5) variants that have potential functional consequences as they ranked at top 5% of the rank score of CADD (Combined Annotation Dependent Depletion), or DANN (deleterious annotation of genetic variants using neural networks) or fitCons (fitness consequence score) ^80–82^.
6) ‘pathogenic’ or ‘likely pathogenic’ variants as annotated in InterVar database (clinical interpretation of missense variants)^83^;

### Statistical analysis

#### Single-variant-based association test

For variants in non-HLA region, SNVs with a MAF >= 0.001 were included in a logistic regression in which NPC disease status was regressed on the genotype of individual SNV. For variants in HLA region, four types of variants were included in analysis: SNVs, four-digits HLA alleles, six-digits HLA alleles, and amino acids. For each variant, two tests were performed for three binary phenotypes: all NPC cases (ALLNPC) vs controls, and NPC cases with family history (FHNPC) vs controls, respectively. Other covariates in the model includes top five genetic principal components and gender. Bonferroni method was applied in multiple testing correction (N=157,024), with significance threshold P value of 3.2 × 10^-7^.

#### Conditional analysis for single-variant-based associations

To test whether the single-variant association from target SNV was independent of the associations of other SNVs, conditional analysis was performed by jointly fitting each of the SNVs to be conditioned as covariate variable in the regression model.

#### SNV-set-based association analysis 1: gene-based test

The gene boundary was defined by the transcription start and end sites. These was no MAF-based filtering for this analysis (singletons were also included). Apart from testing using all SNVs (SNV type 1), we also performed test that only consider coding-affecting SNVs. For each SNV type, we tested 4 algorithms that are available on the “SKAT” package in R:

1) The original SNP-set (Sequence) Kernel Association Test (SKAT)^84^.
2) The original Burden test^85^.
3) SKAT for the combined effect of common and rare variants using the sum test as described previously^86^.
4) Burden test for the combined effect of common and rare variants using the sum test as described previously^86^.

For each gene tested using each SNV type for each phenotype, the result from the algorithm with the most significant P value was remained. Top ten genetic principal components and gender were included in the model as covariates. Bonferroni method was applied in multiple testing correction (N=22,228), with significance threshold P value of 2.3 × 10^-6^.

#### SNV-set-based association analysis 2: pathway-based test

The pathway information was downloaded from Molecular Signature Database, including curated canonical pathways, GO Ontology items and oncogenic signature gene sets^79^. After annotating SNVs to the pathways, we performed pathway-level association analysis to test cumulative effect from both rare and common variants within a pathway on NPC risk. To avoid false positive results driven by very small or very large gene set, we excluded pathways with a size larger than 200 genes or smaller than 5 genes. The same grouping and algorithms applied in the gene-based test were also applied here. Multiple testing correction was performed using the FDR method (N_pathway_=6,204).

#### Meta-analysis for single-variant-based and gene-based associations

The random effect model (REML) was applied to meta-analyze the association between individual variants and NPC using the R package “metafor”^87^. The summation of logits method^88^ was applied to meta-analyze the association between individual genes and NPC using the R package “metap”^89^.

#### Structural modelling of E-selectin

The molecular structure of the E-selectin complexed with glycomimetic antagonist 2-acetamido-2-deoxy-beta-D-glucopyranose (PDB code 4C16) was modelled as previously described^90^. PyMOL with the Adaptive Poisson-Boltzmann Solver plugin was used in the evaluation of the electrostatic surface and the visualization.

#### Assessing cell-type specific gene expression of NPC and other cancer-associated genes using scRNA-seq data of tumor and non-tumor tissues

Susceptibility gene list of NPC, lung, colorectal and gastric cancers included those identified in present study (for NPC only) and those collected from published GWAS studies (**Table S10)**. Expression profiles were derived from multiple scRNA-seq datasets: the NPC tumor dataset (n_sample_=56, n_cell_= 223,593) as described in “NPC samples for Single-cell RNA-seq” section; the lung cancer tumor dataset (n_sample_=19, n_cell_=90,477) downloaded from the GEO database (GSE131907, GSE139555) and collected from a previous pan-cancer study^91^; the gastric cancer tumor dataset (n_sample_=9, n_cell_=28,299) provided by a published study^92^; the colorectal cancer tumor dataset (n_sample_=37, n_cell_=70,015) downloaded from the GEO database (GSE132465, GSE144735 and GSE139555); and the non-cancer tissue dataset consisting of 483,152 cells (for computational consideration, 100,000 cells were randomly selected in downstream analyses) from 10 tissues of 15 donors provided by a published study^37^. The quantification of gene expression and determination of cell types were performed separately for each dataset.

Specifically, for NPC data we first used DoubletFinder to detect doublets^93^ by inspecting abnormal clusters that simultaneously express two or more cell type markers. For each sample, doublets were removed with the expected doublet rate of 0.05. Cells with n_Feature_RNA_ > 500, n_Count_RNA_ > 1,000 and percent.mt < 25% were retained for downstream analyses. Expression Matrix of all cells were converted to a Seurat object and managed by Seurat’s pipeline^94^: normalization with NormalizeData and ScaleData, batch effect correction with Harmony^95^, dimensionality reduction with RunUMAP, clustering with FindNeighbors (k.param = 30) and FindClusters. Clusters were annotated by classic markers as following: NK cells (KLRF1), T cells (CD3D), Plasma cells (MZB1), B cells (MS4A1), Myeloid cells (AIF1), Cancer Associated Fibroblasts (COL1A1), Endothelial cells (VWF) and Epithelial cells (EPCAM). Normalized expression of marker genes for each cell type in each tumor datasets were shown in **Figure S12**.

Lung, gastric, colorectal cancer tissue datasets were merged and analyzed. We first used Scrublet^96^ to remove estimated doublets and SoupX^97^ to remove cell-free mRNA contamination. Cells with n_Feature_RNA_ > 500, n_Count_RNA_ > 1000 and percent.mt < 25% through quality control were kept for downstream analysis. NormalizeData and ScaleData in Seurat was used to normalize data^98^; FindVariableFeatures with n_feature_ = 3000 was used to select hypervariable genes. We used Harmony to correct for batch effect caused by different patients^95^, and used RunUMAP with dims = 1:30 and FindClusters with resolution = 0.3 for dimension reduction and clustering. The clusters were annotated by classic markers of major cell types. Normalized expression of marker genes for each cell type in each tumor datasets were shown in **Figure S12**.

For the non-cancer tissue dataset, cell clusters were determined by the original study^37^.

#### Inference of malignant status of epithelial cells in scRNA-seq data in NPC tumor tissues

Malignant cells have substantially increased copy number variation (CNV), such as large-scale gain or deletions of chromatin. Therefore, we use inferCNV to identify malignant epithelial cell clusters^99^. Comparing with control normal epithelial cells downloaded from GEO database (GSE121600), all epithelial cell clusters identified in NPC scRNA-seq data in our study were considered malignant.

#### Estimate of WES-SNV heritability for NPC

GREML-LDMS (the LD and MAF stratified GREML) approach was applied to estimate the proportion of NPC risk that can be explained by WES SNVs ^100^. In brief, when calculating the total WES SNV-heritability and the proportion explained by variants from HLA and non-HLA regions, SNVs were stratified into four groups based on MAF and regions: MAF ≥ 0.01 in HLA region, MAF<0.01 HLA region, MAF ≥ 0.01 in non-HLA region and MAF < 0.01 in non-HLA region; when further decomposing the WES SNV-heritability in non-HLA region, SNVs were stratified into 28 groups based on LD scores (four bins based on quartiles. LD score was calculated as the sum of R^2^ with variants in a 200kb window) and MAF (7 bins: 0.0005 < MAF <= 0.001, 0.001 < MAF <=0.01, 0.01 < MAF <=0.1, 0.1 < MAF <= 0.2, 0.2 < MAF <=0.3, 0.3 < MAF <=0.4, and 0.4 < MAF <=0.5). For each SNV group, genomic relationship matrix (GRM) was created. In each model (the four components and 28 components ones), GRMs from all groups were then simultaneously fitted in the GREML model for NPC, where the total NPC variation explained by all genetic components and the variation explained by individual genetic component as represented by each GRM can by estimated.

#### Joint model for a comparative analysis to estimate the relative risks by RPL14, SELE, HLA, and non-HLA GWAS loci

In discovery samples, a subset (1,382 cases and 912 controls) of the GD-SYSUCC cohort simultaneously have imputed GWAS array^17^ and WES data available, allowing the comparisons of the relative disease risks conferred by the common variant rs2276868 in *RPL14* and the rare variant *rs5361* in SELE with the polygenic risk from the HLA and other known GWAS loci. In replication samples, the cap-seq designed to target at both published GWAS loci and the candidate loci identified by the presented WES study also enabled this comparison. In brief, for each cohort we applied a joint regression model that simultaneously accounts for genetic effects from different variants: the effects from *RPL14* and *SELE* were presented by the genotypes of rs2276868 (*RPL14*) and rs5361(*SELE*), respectively; the combined genetic effects from published non-HLA GWAS loci and that from HLA loci were represented as two polygenic risk scores (one for non-HLA GWAS loci and one for HLA loci) constructed using odds ratios estimated from original publication or our single-variant analysis using discovery cohorts (**Table S14**).

#### Construction and evaluation of polygenic risk scores (PRS) for NPC

##### 1. Construction of a refined composite PRS (rcPRS) for NPC risk

The rcPRS was calculated for all individuals with non-missing genotypes in the discovery sample (the subset of 1,382 cases and 912 controls with both GWAS array and WES data available^17^ in GD-SYSUCC cohort) and in all replication samples (the capture sequencing allowed the genotyping for constituent variants of combined PRS). The following SNVs were used to create rcPRS to simultaneously account for genetic risk factors identified in our WES analyses and published GWASs^8,17,101^:

1) In WES data, significant variants in single-variant-based association test (including HLA variants identified in conditional analysis).
2) In WES data, leading variants in gene-based test if they:

a. a. were in genes significant in SKAT-gene-based association test.
b. reached P<1×10^-4^ in single-SNV-based association test.
3) In WES data, leading variants in pathway-based test as they:

a. were in a gene of significant pathways and that gene has a P<0.01 in gene-based test.
b. reached P<1×10^-4^ in single-SNV-based association test.
4) Published GWAS loci ^8,17,101^. Only non-HLA GWAS loci was extracted from this source as majority of HLA SNPs identified in published GWAS were in non-coding regions, which are beyond the regions captured by WES and that signals from HLA regions have already been included in source 1.

To avoid duplication due to overlapped genes among pathways, each variant can be used only once. Clumping was performed so that in each cluster of LD partners with R^2^>=0.1 (within 250kb extended windows) only one variant could be remained. PRS was created as the sum of weighted genetic risk from all variants. For each variant, the weighted genetic risk was calculated as the product of allele dosage and the coefficient of variant effect estimated in single-variant-based test. For variants from published GWAS, the coefficient was extracted from original publications ^8,17,101^. Details of the constituent variants were shown in **Table S15**.

##### 2. Construction of GWAS-only PRS (gPRS) for NPC risk

For a subset sample (n=2,294) of the discovery GD-SYSUCC cohort, we produced imputed genotypes for BeadChips array data. In brief, SNP array data was obtained from a previous study^8^. To prepare the data for imputation analysis, we first used Liftover to convert the SNP array data from hg18 to hg19 encoding strand. Next, we performed quality control at the SNP level, removing SNPs with a minor allele frequency (MAF) less than 0.01, genotype missing rate greater than 0.05, and those that failed the Hardy-Weinberg equilibrium test (HWE) with a P-value less than 10^-5^. A total of 463,270 SNPs passed quality control and were retained for further analysis. For the quality-controlled genotype data, we used SHAPEIT2 with default parameters to estimate haplotypes of the study samples^102^. We then used IMPUTE2 with default parameters and the haplotype data from the 1000 Genomes Project to impute the missing genotype of each individual’s haplotype^103^. After imputation, we removed SNPs with an INFO value less than 0.8 and set loci with genotype probability (GP) values less than 0.9 to missing. Ultimately, we retained 6,095,964 SNPs for analysis. This allows the construction of a previous published PRS using NPC-associated loci reported by GWAS^10^, following the instruction from the original study^10^. To note, for three SNPs (rs2106123, rs6475604, rs9507124) not genotyped and not imputed in GD-SYSUCC, we used three surrogate SNPs in complete LD (R^2^=1 in Chinese Southern Han population in 1000 genome phase 3 dataset) with them (rs6774494 for rs2106123, rs1412829 for rs6475604, and rs9510790 for rs9507124) in the model.

#### Construction of composite scores for pathway and EBV activity

We applied the AddModuleScore function in R package Seurat ^98^ to profile each pathway from the REACTOME and the GO Ontology database in each sample, where bulk RNA-seq or scRNA-seq data was available. Based on AddModuleScore, the pathway score is obtained by subtracting the average expression of the corresponding control gene set from the average expression of the target gene set. For samples with scRNA-seq, we took each patient as a datapoint, meaning that the average expression of a given gene across all epithelial cells from the patient was treated as the expression of that gene for the patient. We additionally created composite scores for EBV activity. In brief, we downloaded the EBV gene list from the EBV reference sequence (Akata; https://github.com/flemingtonlab/public/tree/master/annotation) and mapped the scRNA-seq data of EBV-positive malignant epithelial cells onto them. We then used the annotated EBV gene data to construct an EBV-composite score for each NPC patient with scRNA-seq data derived from EBV-positive epithelial cells available, following the same steps as described above.

#### The correlation between the pathway composite scores and RPL14 expression

Pearson correlations between *RPL14* expression and composite scores were tested using the cor.test function in R. Multiple testing correction was performed using the FDR method. The Pearson correlations of individual EBV genes and *RPL14* were shown in **Table S17.**

### Functional characterization of NPC-associated loci

#### Cell culture

Human NPC cell lines S26, CNE2-EBV, and HONE1-EBV were kindly gifted by Professor Mu-Sheng Zeng at the SYSUCC. Human NPC cell lines HK-1 and C666-1 were kindly gifted by Professor KW Lo at the Chinese University of Hong Kong. Human embryonic kidney cell 293T cell was purchased from Cell Bank of Type Culture Collection of Chinese Academy of Sciences, Chinese Academy of Sciences. All above cells were cultivated in DMEM medium containing 10% fetal bovine serum (FBS, Gibco, NY, USA) and 1% penicillin–streptomycin solution (FBS, Gibco, NY, USA). Human umbilical vein endothelial cells (HUVECs) were isolated and cultured in ECM medium (ScienCell, CA, USA) supplemented with 15% FBS and 0.03 mg/mL of endothelial cell growth supplement (ECGS, ScienCell, CA, USA). All cells were cultured at 37℃ under a humidified atmosphere with 5% CO2. For all cell lines, mycoplasma was routinely determined using PCR with specific primers and no contamination was detected (**Table S18**).

#### Cell proliferation

For cell growth curves, a total of 1×10^5^ NPC cells was plated in a 12-well plate with three replicates. The number of cells was measured by a cell counter with trypan blue staining on 24, 48 and 72h. For colony formation assays, NPC cells were seeded at a density of 3000 cells per replicate into 6-well plate for 7-10 days. Then, colonies were fixed with 4% paraformaldehyde for 30min at room temperature. After stained with crystal violet for 10min, cells were photographed with the Bio-Rad ChemiDoc Touch (Hercules, CA, USA).

#### Cell migration

Transwell assay was performed as described previously^104^. Briefly, NPC cells in serum-free medium were plated at a density of 6×10^4^ cells into the transwell chamber (8 µm pores, Corning, NY, USA), which then placed at 24-well plates containing 600 µL DMEM suppled with 10% FBS. After 16-24h, cells on the membrane were fixed with 4% paraformaldehyde for 30min and stained with trypan blue solution for 15min at room temperature. Then the cells were imaged under a microscope.

#### Luciferase reporter assay

Luciferase reporter assay was performed as described previously^105^. Briefly, the promoter region of RPL14 containing the rs2276868-C or -T was cloned into a luciferase reporter pGL3-basic plasmid. Afterwards, 293T cells were co-transfected with 200 ng of pGL3-RPL14-C or -T constructs and 10 ng of pRL (Renilla luciferase) plasmid together with respective siRNAs or overexpression plasmids. After 48h, cells were harvested to measure the luciferase activity through a Dual-Luciferase Assay Kit (Promega, USA) according to the manufacturer’s instructions.

#### Immunofluorescence staining (IF) and immunohistochemistry (IHC)

Paraffin slides of xenograft tumors were deparaffinized with xylene and rehydrated through a gradient ethanol series. After microwave antigen retrieval in sodium citrate solution (pH 6), slides were incubated in 0.3% hydrogen peroxide for 15 min at room temperature to quench the endogenous peroxidase activity. Afterwards, the samples were incubated with the primary anti-Ki-67 (AB_393778, BD Pharmingen™, USA) or CD34 (GB13013, Servicebio, China) antibodies overnight at 4 °C. Next day, for IF, Alexa Fluor-conjugated secondary antibody

(Thermo Fisher Scientific, USA) was used for 1 h at room temperature, followed by counterstaining with antifade mounting medium and 4′,6-diamidino-2-phenylindole (DAPI) for 10 min. Images were then captured with a confocal laser scanning microscope (Carl Zeiss, Microscope 880, Germany). For IHC, after exposure to secondary antibodies at room temperature for 1h, chromogenic detection was performed using 0.05% 3,30-diaminobenzidine (Dako, Denmark). Hematoxylin was applied for counterstaining. Two independent pathologists from SYSUCC evaluated staining intensity. Afterwards, IHC scores were then calculated, combining staining intensity and the proportion of stained cells.

#### Manipulating gene expression using siRNA and lentivirus

For transient knockdown of target genes, siRNAs specific against RPL14 or NKRF were transfected into cells using Lipofectamine RNAiMAX (Invitrogen, USA) according to the manufacturer’s protocol. After 48-72 hours, cells were harvested to detect the knockdown efficiency of target genes through RT-qPCR as described below. Full length cDNA of NKRF was cloned into the pcDNA3.1 vectors. RPL14, SELE-WT and MUT were cloned into the pCDH-puro lentiviral vectors. For lentivirus preparation, 293T cells were transfected with above lentivirus plasmids or control vector using Lipofectamine 2000 (Invitrogen, USA) according to the manufacture’s procedures. Afterwards, the supernatant media containing lentivirus were collected two times at 24 and 48h post-transfection to infect NPC cell lines, following selection with puromycin (2 μg/mL) at least for one week.

#### RNA isolation, RT-qPCR and ChIP PCR

Total RNA was isolated using Trizol reagent (Invitrogen, USA) following the manufacturer’s protocols. After determination of RNA concentration using a Nanodrop Spectrophotometer (Thermo Fisher Scientific, USA), cDNA was synthesized using oligo (dT) primers and M-MLV Reverse Transcriptase (Promega, USA) according to the manufacturer’s procedures. RT-qPCR was performed with SYBR Premix Ex Taq kit (Takara, Japan) on a LightCycler 96 (Bio-rad, USA). GAPDH or 18S RNA was used as an internal control. The primer sequences are shown in **Table S18**. For ChIP assay, S26 cells transfected with Flag-NKRF plasmids or control vectors were grown in complete DMEM media to 80%–90% confluency. The media were removed and replaced with media containing 1% formaldehyde and crosslinked for 10 min at 37°C, followed by SimpleChIP® Plus Enzymatic Chromatin IP Kit (Magnetic Beads; CST, USA) according to the manufacturer’s instructions. Afterwards, PCR or qPCR was performed with specific detection primers following the standard procedures (**Table S18**).

#### EBV preparation and infection

AKATA cells carrying EBV recombinant viruses were treated with 0.75 (v/v) of goat anti-human immunoglobulin G IgG (H0111-6, Tianfun Xinqu Zhenglong Biochem, China) for 6h at 37℃ to induce the viral productive cycle. Virus supernatants were harvested three days post-induction, filtered through a 0.45 µm filter and concentrated at 20000g for 3h. Afterwards, virus concentrate was added into NPC cells and concentrated at 2000g in 37℃ for 1h. Cell supernatants were removed and replaced with normal media 6-8h post-infection. The infection efficiency was measured using flow cytometry analysis of GFP-positive cells.

#### Western blotting

NPC cells were lysed in ice-cold cell lysis buffer (CST, USA) containing 1X protease inhibitors (Beyotime, China). Lysate was cleared by centrifugation at 14,000g for 10 min at 4°C and boiled in 1X LDS loading buffer at 100℃ for 10 min. Proteins were separated by SDS-PAGE and transferred to 0.2 μm polyvinylidene difluoride (PVDF) membrane (Merck Millipore, USA). Membranes were blocked with 5% bovine serum albumin (BSA; Sangon Biotech, China) in tris-buffered saline and Tween 20 (TBST) at room temperature for 1h. Afterwards, the membranes were incubated with target-specific antibodies in blocking buffer at 4°C overnight. Next day, horseradish peroxidase–conjugated (HRP) secondary antibodies were used at room temperature for 1h. Proteins were then detected with Fdbio-Dura ECL kit (Fdbio science, China) and quantified by Bio-Rad ChemiDoc Touch (Hercules, USA). Primary antibodies were commercially available, including RPL14 and LMP1 (ab181200 and ab78113, Abcam, USA), SELE (20894-1-AP, Proteintech, China), Flag (F1804, Sigma-Aldrich, USA), BZLF1(sc53904, Santa Cruz, USA) and ACTIN (AC004, Abclonal, China).

#### In vivo tumor xenograft

Six-week-old male BALB/c nude mice (Beijing Vital River Laboratory Animal Technology, China) were grown in the Specific Pathogen Free (SPF) environment. For subcutaneous xenograft assay, 5×10^6^ CNE2-EBV cells or 2×10^6^ S26 cells co-culturing with HUVEC cells (S26: HUVEC = 10:1) were suspended in 100µL ice-old PBS mixed with Matrigel (0.20 v/v, Corning Incorporated, USA) and subcutaneously injected to the two flanks of the mice. Macroscopic observation and tumor volume measurement using a caliper were performed every 3 days. After 15 days, all mice were sacrificed, and tumor tissues were carefully dissected and weighed. Tumor volume was calculated following the formula: tumor volume (mm3) = length (mm) × (width (mm)) 2 /2. For tumor lymphatic metastasis assay, 1×10^6^ S26 cells co-culturing with HUVEC cells (S26: HUVEC = 10:1) were injected into the foot pads of the node mice. After one month, the lymph nodes were harvested, volumed and paraffin embedded further HE staining. For animal studies *in vivo*, all experiments were performed in strict accordance with the instructions approved by the Institutional Animal Care and Use Committee of Sun Yat-sen University.

#### Tube formation assays

For tube formation assays, HUVEC cells stably expressing SELE wild-type (SELE-WT), the S149R mutant or control vectors were resuspended in the ECM medium. Then the cells were seeded into 96-well plates pre-coated with growth-factor-reduced Matrigel (Corning Incorporated, USA) and incubated at 37°C for 5 hours. The formation of capillary-like structures was subsequently observed using a microscope. Quantitative analysis was conducted by counting the number of tube networks formed across the entire field, serving as an indicator of *in vitro* angiogenesis capability.

