## Supplementary figures for "Large-Scale Whole-Exome Sequencing Association Study Implicates Genetic Effects on Viral Oncogenesis and Tumor Microenvironment in Nasopharyngeal Carcinoma"

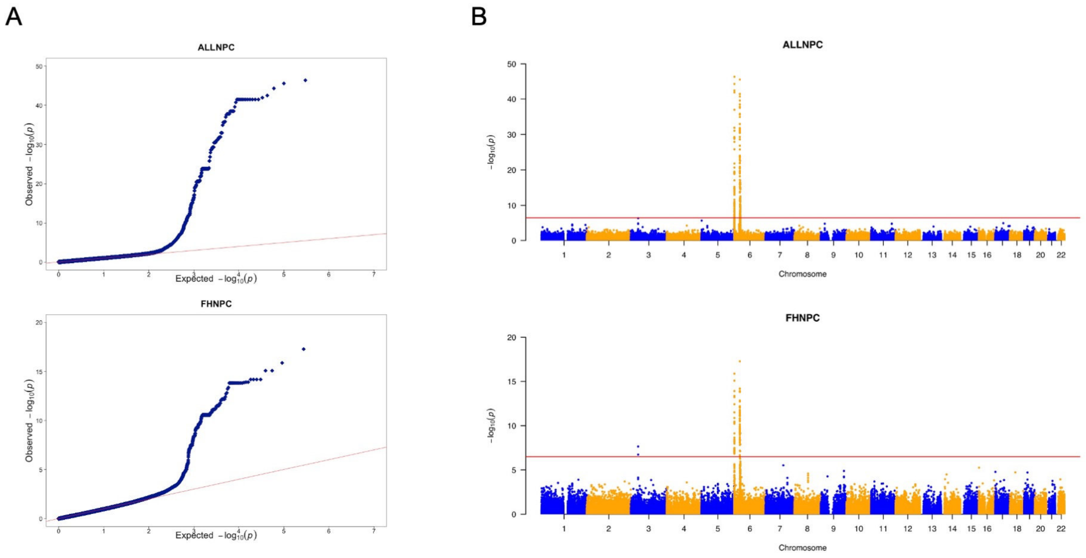


**Figure S1. QQ and Manhattan plots of single-variant-based association test.** ALLNPC: all NPC cases. FHNPC: NPC cases with a family history of NPC. **A.** QQ plots. **B.** Manhattan lots. Red line: significant threshold based on Bonferroni's method ($P < 3.2 \times 10^{-7}$).

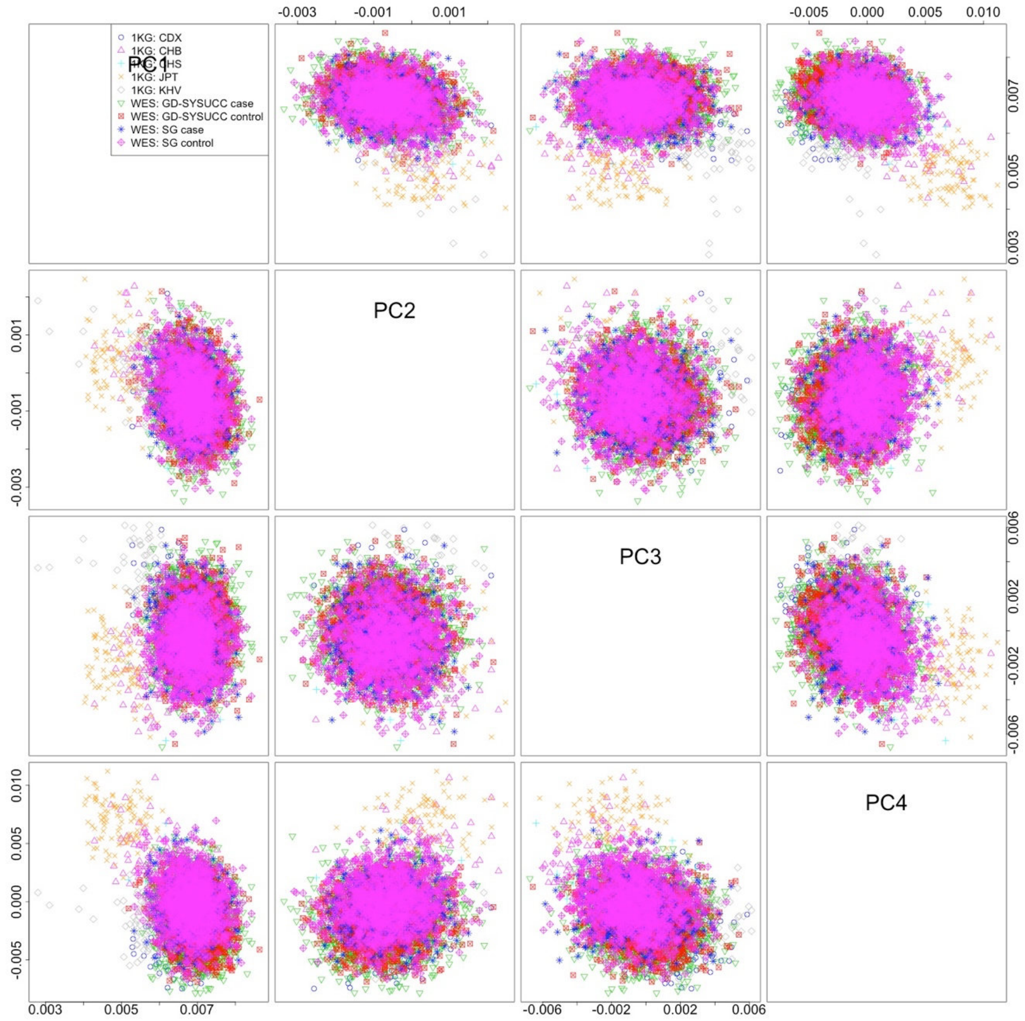


**Figure S2. Top four genetic principal components of the discovery samples and the East Asia samples in 1000 genome reference data (phase 3).** 1KG: population samples in 1000 genome reference data (phase 3); CDX: Chinese Dai in Xishuangbanna, China; CHB: Han Chinese in Beijing, China; CHS: Han Chinese South, China, China; JPT: Japanese in Tokyo, Japan; KHV: Kinh in Ho Chi Minh City, Vietnam. PC: principal component.

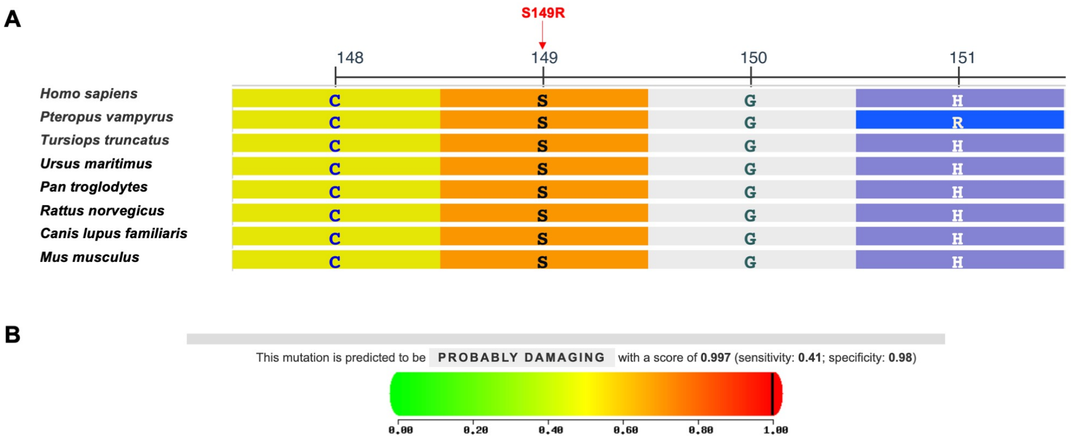


**Figure S3. Functional implication of rare variants in SELE to NPC. A.** Comparative genomic analysis of amino acid status across multiple species at position 149 and adjacent regions in SELE using NCBI Multiple Sequence Alignment Viewer. **B.** The rs5361 variant is predicted as “probably damaging” according to Polyphen-2.

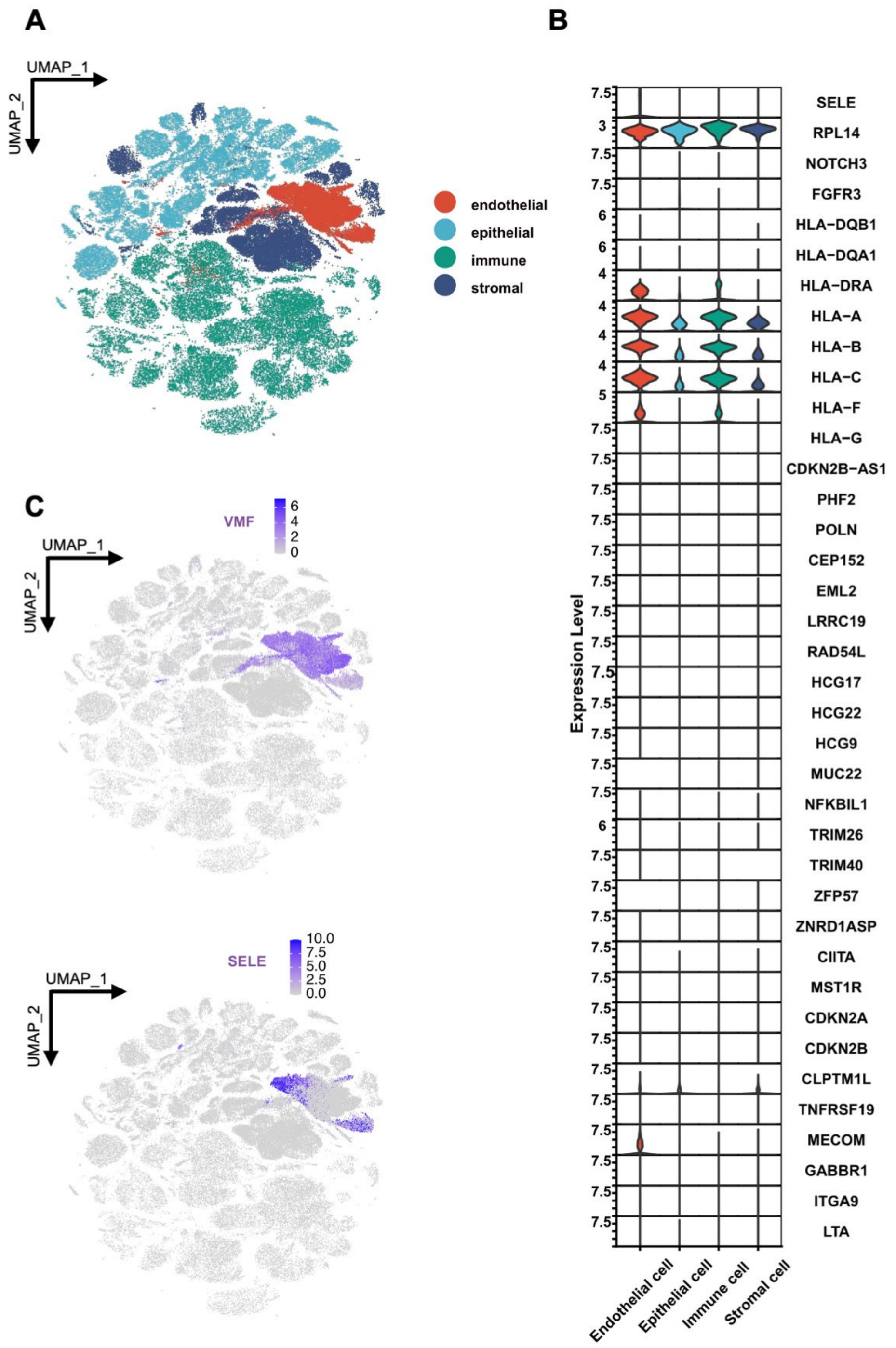


**Figure S4. Expression patterns of novel and known NPC-associated genes across diverse cell types in non-tumor tissues.** Single-cell transcriptomic analyses of 100,000 cells from non-tumor tissues of 15 individuals without cancer. **A**. UMAP plot of 100,000 single cells grouped into four major cell clusters. **B**. Violin plot of normalized expression of NPC-associated genes in major cell clusters. **C**. The expression of the maker gene (VWF) for endothelial cells alongside the novel NPC-associated gene *SELE*.

**A**

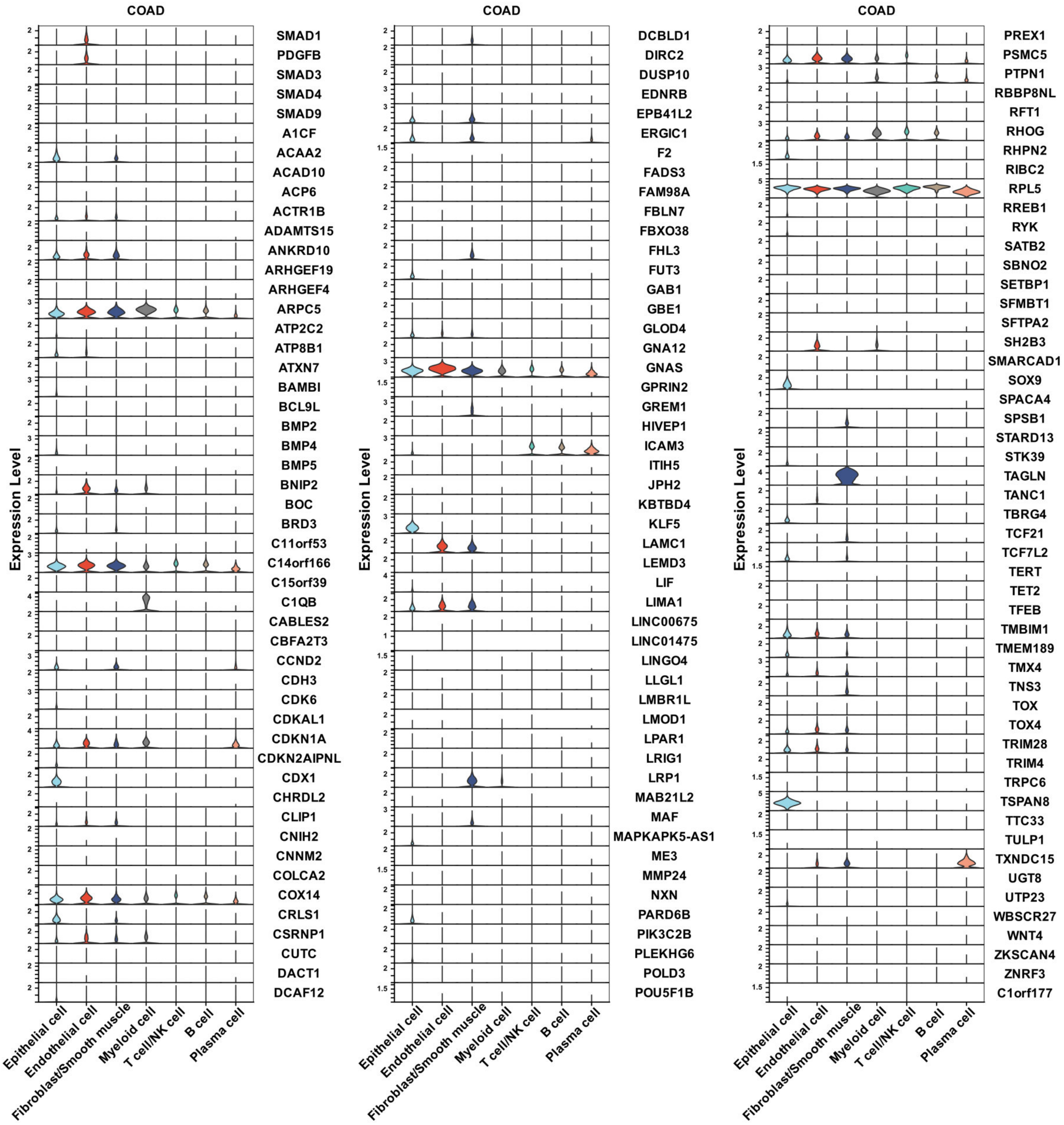
COAD
Expression Level
SMAD1
PDGFB
SMAD3
SMAD4
SMAD9
A1CF
ACAA2
ACAD10
ACP6
ACTR1B
ADAMTS15
ANKRD10
ARHGEF19
ARHGEF4
ARPC5
ATP2C2
ATP8B1
ATXN7
BAMBI
BCL9L
BMP2
BMP4
BMP5
BNIP2
BOC
BRD3
C11orf53
C14orf166
C15orf39
C1QB
CABLES2
CBFA2T3
CCND2
CDH3
CDK6
CDKAL1
CDKN1A
CDKN2AIPNL
CDX1
CHRDL2
CLIP1
CNIH2
CNNM2
COLCA2
COX14
CRLS1
CSRNP1
CUTC
DACT1
DCAF12
COAD
Expression Level
DCBLD1
DIRC2
DUSP10
EDNRB
EPB41L2
ERGIC1
F2
FADS3
FAM98A
FBLN7
FBXO38
FHL3
FUT3
GAB1
GBE1
GLOD4
GNA12
GNAS
GPRIN2
GREM1
HIVEP1
ICAM3
ITIH5
JPH2
KBTBD4
KLF5
LAMC1
LEMD3
LIF
LIMA1
LINC00675
LINC01475
LINGO4
LLGL1
LMBR1L
LMOD1
LPAR1
LRIG1
LRP1
MAB21L2
MAF
MAPKAPK5-AS1
ME3
MMP24
NXN
PARD6B
PIK3C2B
PLEKHG6
POLD3
POU5F1B
COAD
Expression Level
PREX1
PSMC5
PTPN1
RBBP8NL
RFT1
RHOG
RHPN2
RIBC2
RPL5
RREB1
RYK
SATB2
SBNO2
SETBP1
SFMBT1
SFTPA2
SH2B3
SMARCAD1
SOX9
SPACA4
SPSB1
STARD13
STK39
TAGLN
TANC1
TBRG4
TCF21
TCF7L2
TERT
TET2
TFEB
TMBIM1
TMEM189
TMX4
TNS3
TOX
TOX4
TRIM28
TRIM4
TRPC6
TSPAN8
TTC33
TULP1
TXNDC15
UGT8
UTP23
WBSCR27
WNT4
ZKSCAN4
ZNRF3
C1orf177
Epithelial cell
Endothelial cell
Fibroblast/Smooth muscle
Myeloid cell
T cell/NK cell
B cell
Plasma cell

**B**

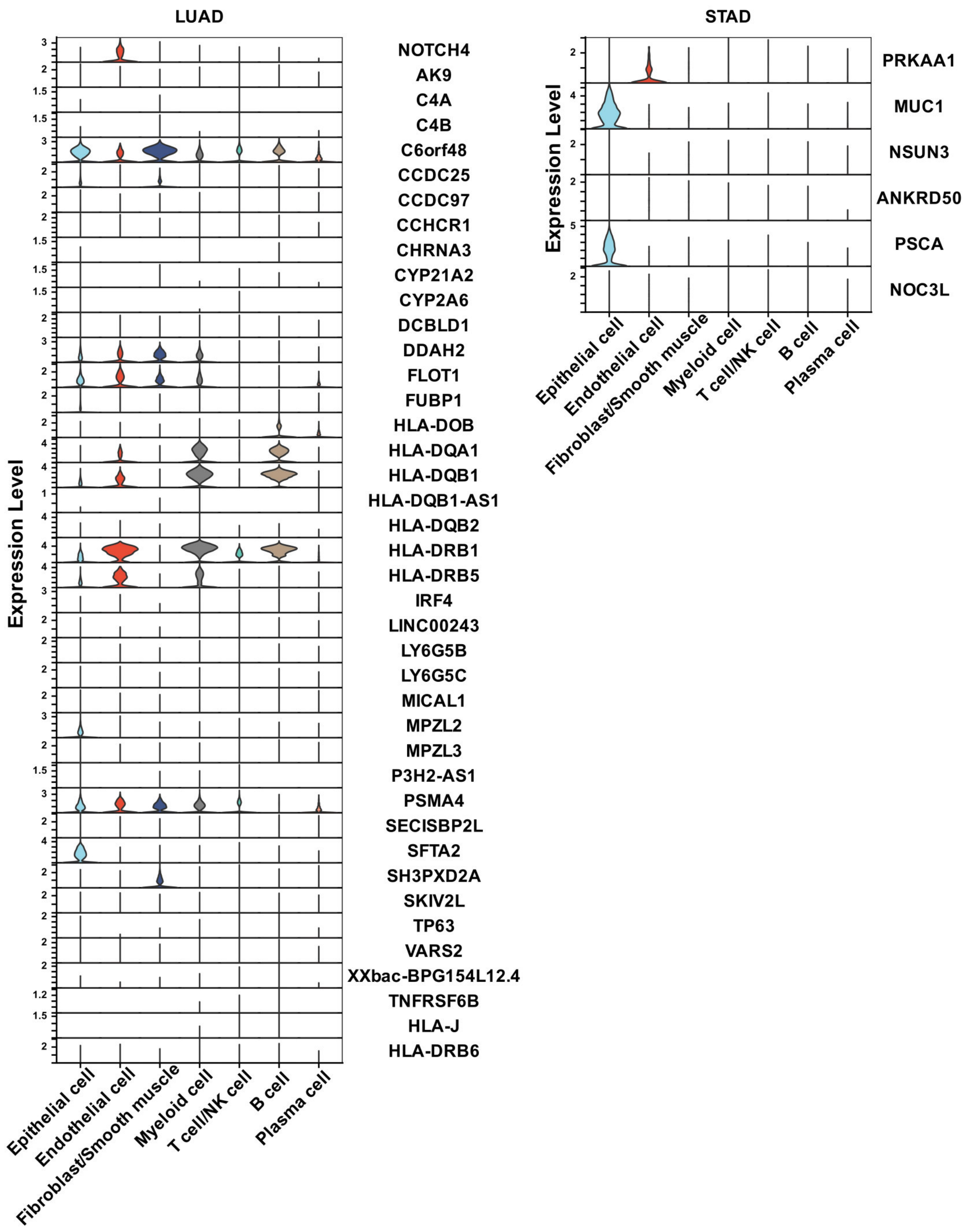


**Figure S5. Cancer-associated genes were expressed in diverse cell types in tumor tissues.** **A.** Colon adenocarcinoma (COAD)-associated genes' expression pattern in colorectal tumor tissues. **B.** Lung adenocarcinoma (LUAD) and Stomach adenocarcinoma (STAD)- associated genes' expression patterns in lung and gastric tumor tissues, respectively. Associated gene list was collected from published studies (see Methods). Expression level: normalized mRNA expression (see Methods).

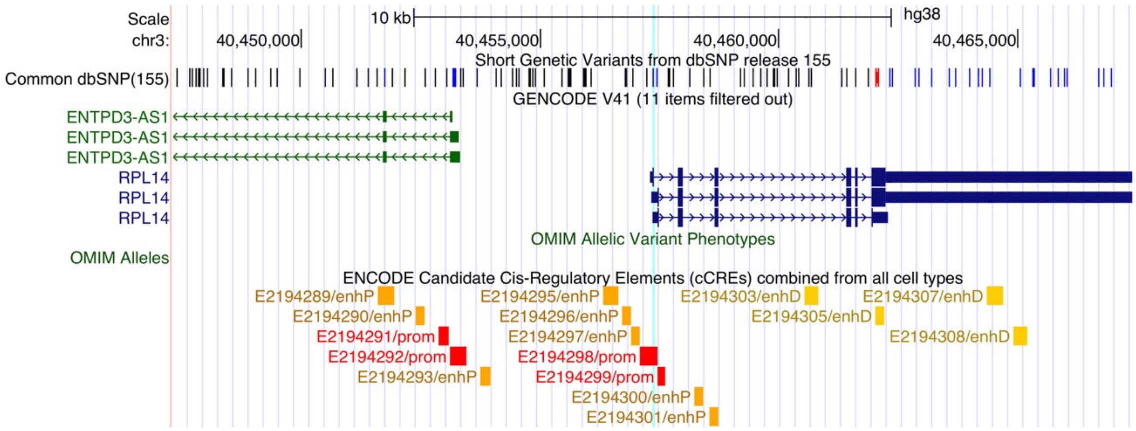


**Figure S6. Genomic annotation of the NPC-associated SNP rs2276868.** Light blue vertical line: The position of the target SNP rs2276868. The overlap between the light blue vertical line and a *cis*-regulatory element (E2194298, promoter) reported by the ENCODE consortium as well as the 5’UTR region of *RPL14* implicated that rs2276868 has the potential to regulate the gene expression of *RPL14* in *cis*. The figure was produced by UCSC genome browser (https://genome.ucsc.edu/).

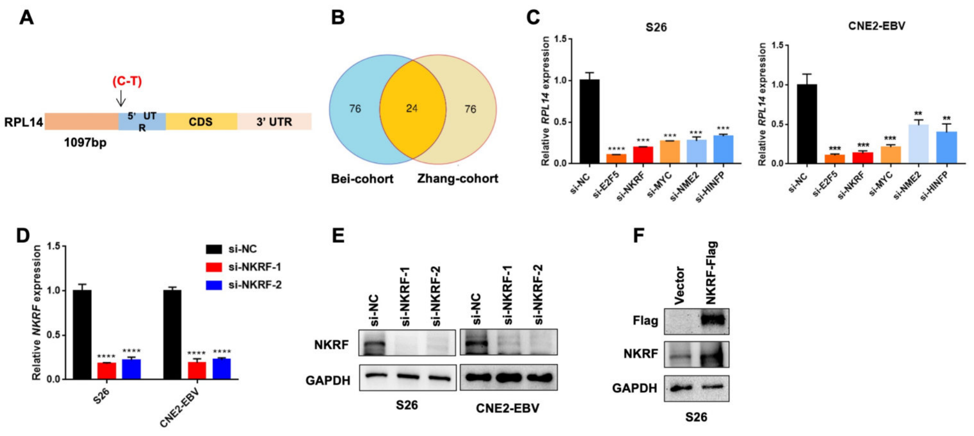


**Figure S7. Candidate transcription factors of *RPL14*. A.** Schematic diagram presented the DNA fragments of RPL14. **B.** Venn diagram showed the overlap transcription factors correlated with *RPL14* expression in two independent NPC cohorts. **C.** S26 and CNE2-EBV cells were transfected with indicated siRNAs targeting candidate transcription factors of *RPL14* or control siRNA. RT-qPCR was performed to detect the mRNA expression of *RPL14*. **D-E.** RT-qPCR (D) and western blotting (E) results showed the knockdown efficiency of *NKRF* in S26 and CNE2-EBV cells transfected with *NKRF* siRNAs or control siRNA. **F.** S26 cells were transfected with NKRF overexpression plasmids or control vector. Western blot was performed to evaluate the protein levels of NKRF. GAPDH was used as internal control. *$P<0.05$, **$P<0.01$, ***$P<0.001$, ****$P<0.0001$.

**A**

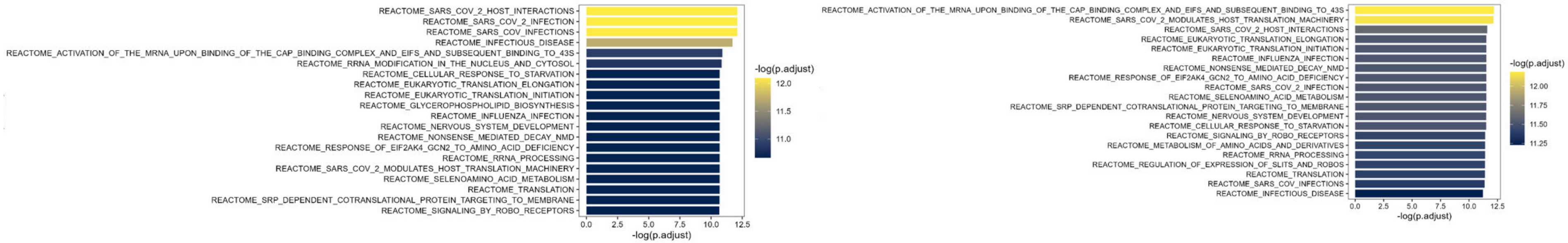


**B**

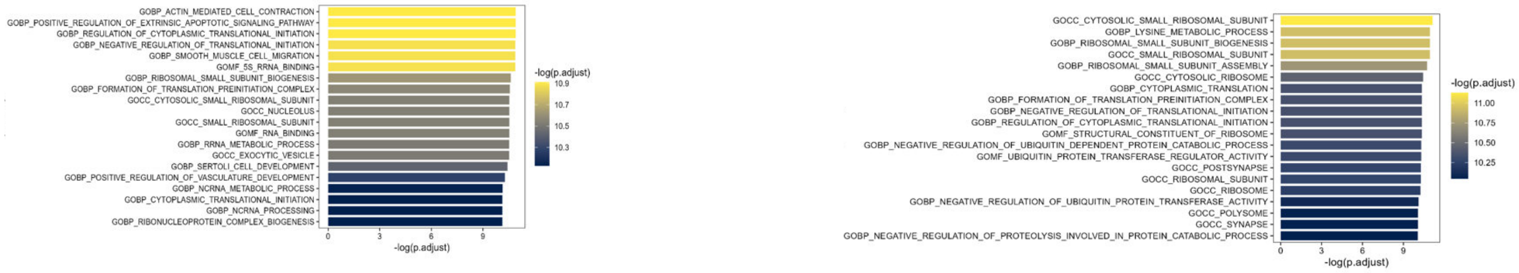


**Figure S8. The top 20 pathways significantly correlated with the expression of *RPL14* in tumor tissues (Bulk RNA seq data). A.** REACTOME pathways. **B.** GO ontology pathways. Left: the results from the Bei-lab dataset ($n_{sample}$=93). Right: the results from the Zhang-lab dataset($n_{sample}$=113). -log(p.adjust): minus log-transformation of adjusted P value using the FDR method.

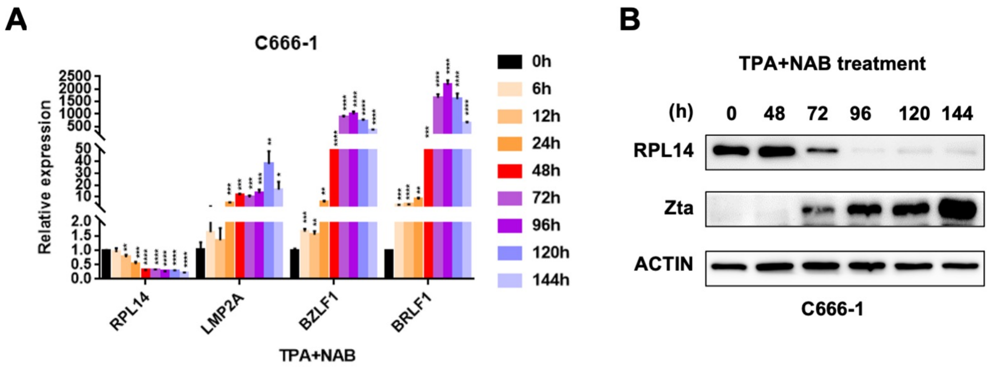


**Figure S9. RPL14 is downregulated with EBV lytic in NPC cells.** C666-1 cells were treated with TPA and NAB to induce EBV lytic. RT-qPCR (**A**) and western blotting (**B**) assays were performed to detect the mRNA and protein expression of RPL14 and EBV genes at different timepoints of EBV lytic. **P*<0.05, ***P*<0.01, ****P*<0.001, *****P*<0.0001.

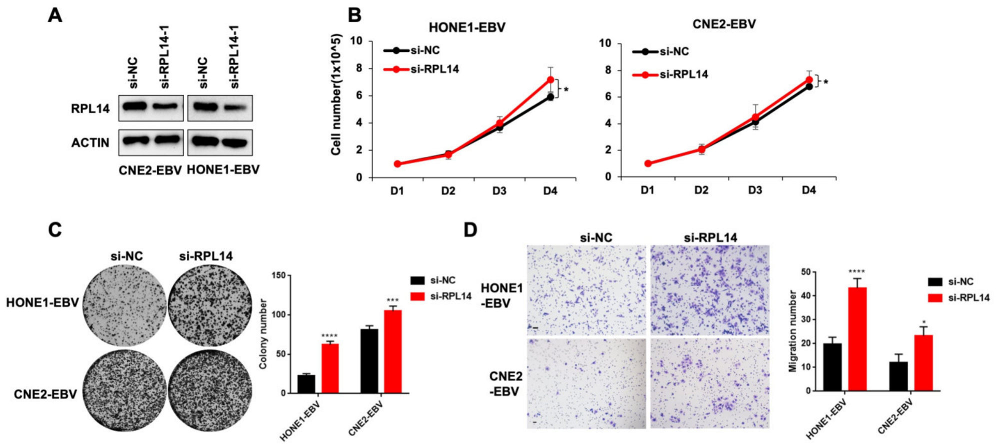


**Figure S10. Knockdown of RPL14 promoted cell proliferation and migration of NPC cells.** **A.** HONE1-EBV and CNE2-EBV cells were transfected siRNAs targeting RPL14 or control siRNA. Western blotting results demonstrated the knockdown efficiency. **B.** Cell growth curves were measured with cells described in A. **C.** Colony formation assays were performed with cells described in A. The statistical data were presented at the right. **D.** Transwell assays were performed with cell described in A and the corresponding statistical analysis were demonstrated at the right. Scale bar, 100 μm. **P*<0.05, ***P*<0.01, ****P*<0.001, *****P*<0.0001.

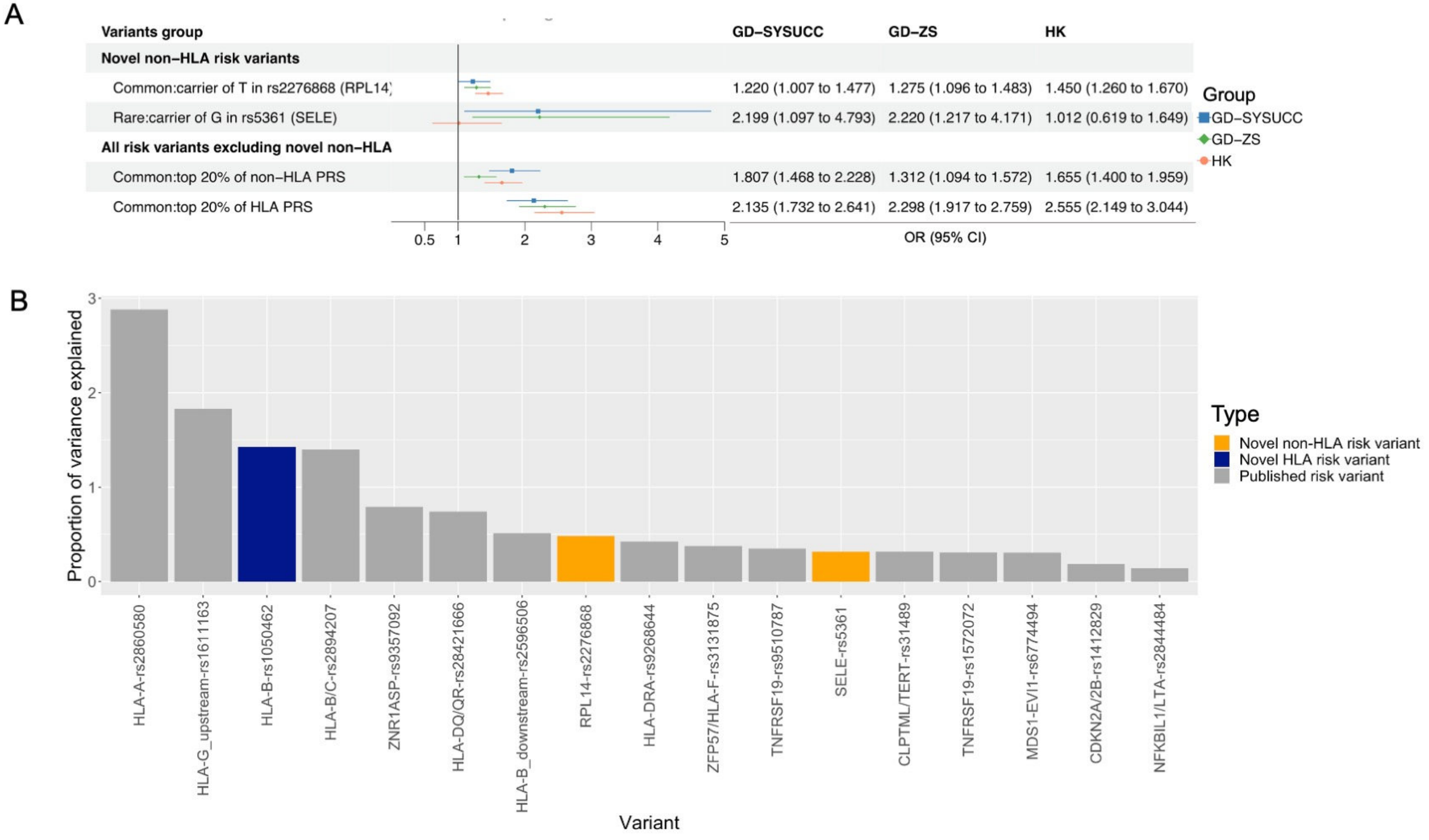


**Figure S11. Dissecting genetic risk for NPC contributed by the novel and previously known loci**. **A**. Detailed odds ratio representation for NPC risk attributed from genetic effects of different variant categories in sample groups. **B**. bar chart visualization of the disease risk broken down by variant types. Orange: novel NPC-associated variants from the non-HLA region, blue: novel NPC-associated variant within HLA region, grey: previously known NPC-associated variants through GWAS.

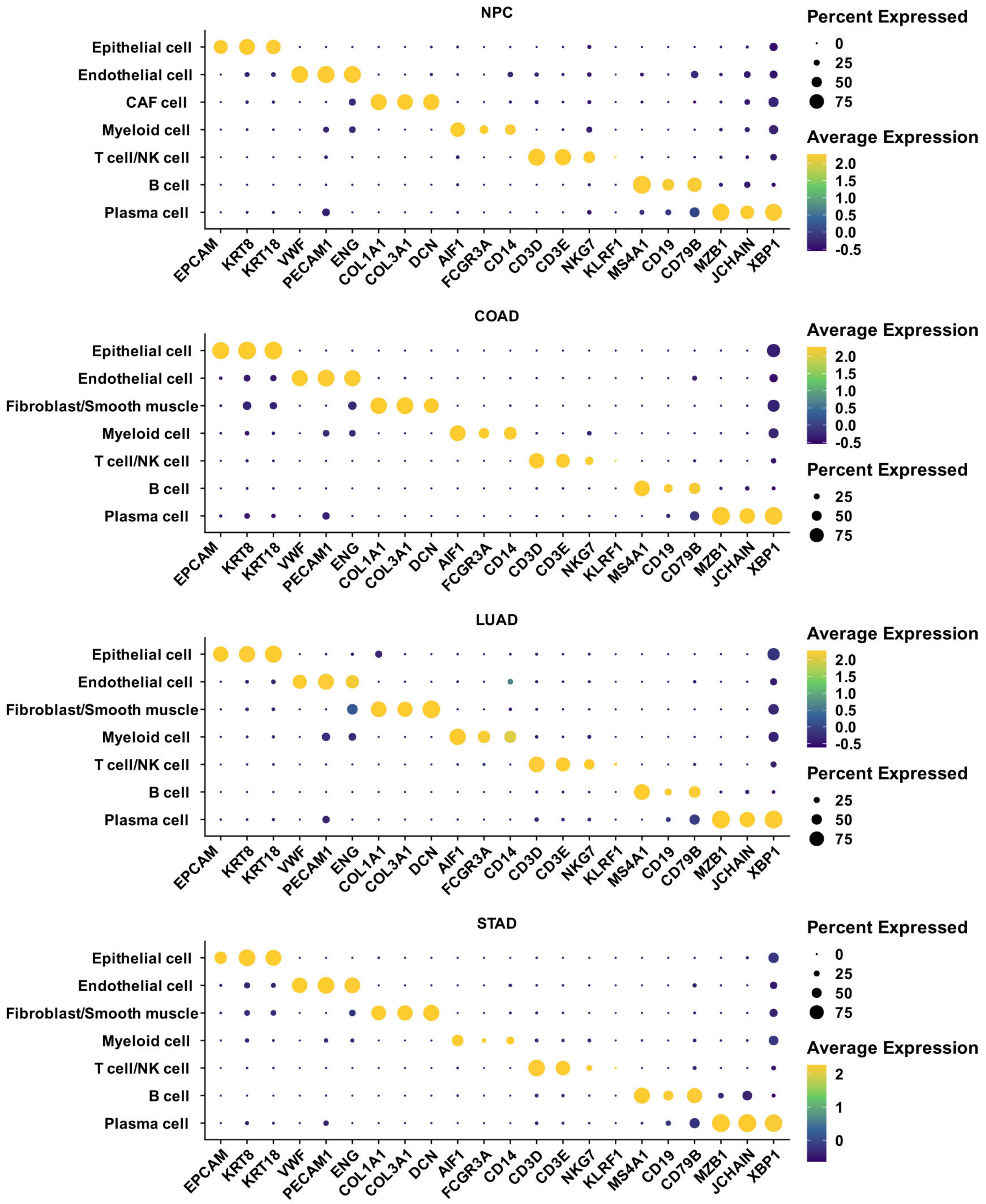


**Figure S12. Normalized expression of marker genes in identified cell types using scRNA-seq data for tumor tissues.** NPC: nasopharyngeal carcinoma, COAD: colon adenocarcinoma, LUAD: Lung adenocarcinoma, STAD: stomach adenocarcinoma. The genes listed were specific marker genes for the major cell types listed in Y axis. Percent expressed: percentage of cells that express a certain marker gene among cells annotated to a given cell type. Average expression: the mean of the normalized expression level of a given marker gene among cells annotated to a given cell type.
